# Antidepressant desvenlafaxine identified in wastewater promotes transformation and antibiotic resistance risk in *Acinetobacter baylyi* via metabolic adaptations

**DOI:** 10.64898/2026.06.02.26353323

**Authors:** Najmuj Sakib, Liezel Mari M. Abaya, Brandon Ruddell, Diana S. Aga, Adina Howe, Laura Jarboe

## Abstract

Wastewater treatment plants (WWTPs) are known reservoirs of antibiotic resistance genes (ARGs). Non-antibiotic compounds such as antidepressants may further promote ARG acquisition through horizontal gene transfer (HGT). Desvenlafaxine, a serotonin-norepinephrine reuptake inhibitor (SNRI) listed on the EU Surface Water Watch Lists, is among the most frequently detected antidepressants in WWTP effluents, yet its role in HGT has not been examined. Here, we detected desvenlafaxine at the highest concentrations among four antidepressants monitored across three municipal WWTPs in western New York. Using *Acinetobacter baylyi* ADP1 as a model recipient in natural transformation assays (n = 6), we found that desvenlafaxine significantly increased transformation frequency at 10 mg/L (1.74 ± 0.33-fold) and 50 mg/L (1.49 ± 0.19-fold; *P_adj_* < 0.05). Effects were independent of reactive oxygen species or membrane permeability stress, consistent with its very low toxicity (IC_20_ ∼1353 mg/L). Instead, desvenlafaxine induced dose-dependent increases in membrane fluidity and shifts to less negative zeta potentials, suggesting that electrostatic interactions between its cationic amine group and the negatively charged membrane reduce surface repulsion and facilitate plasmid proximity during uptake. Non-targeted proteomics revealed a biphasic response: at 10 mg/L, competence-associated proteins (PilB, ComM) were upregulated and STRING analysis identified networks linked to membrane transport, transcriptional regulation, and envelope remodeling, while no connected network was recovered at 50 mg/L. Electron microscopy confirmed higher pili frequency at both doses. Together, these findings reveal an overlooked role of this non-antibiotic pharmaceutical in promoting ARG spread from wastewater environments.

**Importance:** The spread of antibiotic resistance poses a serious and escalating threat to human health worldwide. While antibiotic use is widely recognized as a key driver, non-antibiotic pharmaceuticals released into the environment through wastewater have received far less attention. Antidepressants are among the most frequently detected drugs in treated wastewater effluents, yet their potential to promote antibiotic resistance transfer in bacteria remains poorly understood. This study demonstrates that desvenlafaxine, one of the most abundant antidepressants found in municipal wastewater, increases the uptake of antibiotic resistance genes in environmental bacteria and identifies the bacterial cell-surface changes that enable this.

## Introduction

Selective pressures towards bacterial antibiotic resistance can arise from both antibiotic and non-antibiotic compounds. Most of these compounds, used in areas like households and the pharmaceutical industry, eventually end up in sewage. The microbial communities in urban wastewater treatment plants (WWTPs) are therefore continuously exposed to a diverse array of stressors. High bacterial cell densities, along with these various stresses, can enrich for resistant populations and facilitate horizontal gene transfer (HGT) (1, 2). Many environmental and opportunistic pathogens, including species frequently detected in WWTPs, use transformation as a primary route of HGT (3, 4). DNA released from dead, damaged, or secreting bacterial cells into the extracellular environment (exDNA) is available for transformation into live cells. Transformation can further stabilize incoming DNA into a new cell as episomal plasmids or integrate it into the chromosome (5). As a result, even if bacteria that carry antibiotic-resistant genes (ARGs) are eliminated, transformation may still occur. More than 80 bacterial species are now known to be naturally transformable, with this number steadily increasing as more are studied (3, 6). Although WWTPs remove many contaminants before discharge, conventional treatment technologies are unable to fully eliminate exDNA and trace pollutants (7–9). Several studies indicates that non-antibiotic pharmaceuticals can modulate bacterial physiology in ways that increase transformation frequency, alter membrane properties, or induce competence-associated structures, which raises concerns that such compounds could indirectly contribute to ARG spread even when at sub-inhibitory concentrations (10–15).

Antidepressants are a particularly relevant class of non-antibiotic pharmaceuticals in this context. After ingestion, a significant fraction of these drugs is excreted unchanged or as active metabolites in urine and feces, entering the sewage system directly. They are widely prescribed at steadily increasing rates, and are frequently detected in influent, effluent, and receiving waters downstream of WWTPs (2, 16). Several compounds in this class have been found to increase HGT rates by creating reactive oxygen species (ROS), increasing membrane permeability, and modulating stress and competence pathways (17–19). Venlafaxine and its active metabolite, desvenlafaxine, are notable for their prevalence and regulatory significance in wastewater pollution (20). In municipal WWTP effluents, they are generally detected at tens to several hundred ng L⁻¹, with some plants approaching ∼0.5–0.7 µg L⁻¹ (20). In contrast, hospital wastewater can contain them at µg L⁻¹ levels, with a maximum reported at 10–12 µg L⁻¹ in single hospital effluents (21). However, even when dissolved concentrations in surface waters remain low, multiple field and laboratory studies show that these antidepressants can bioaccumulate in aquatic organisms, which leads to internal concentrations that exceed those in bulk water (22). A 2018–2023 monitoring campaign across 67 Laurentian Great Lakes Basin sites detected peak surface water levels of ∼3,114 ng/L desvenlafaxine and 700 ng/L venlafaxine in 2022, rising notably after COVID-19 (23). In Europe, the European Commission placed both compounds on the Union-wide surface-water Watch List in 2020 and retained them in the 2022 update to address gaps in occurrence, fate, and risk (24, 25).

Desvenlafaxine itself is a second-generation serotonin–norepinephrine reuptake inhibitor (SNRI) approved (as Pristiq) for major depressive disorder in the United States in 2008 (26). It is one of the most frequently detected compounds in surface waters impacted by WWTP discharges and has been highlighted in regulatory and monitoring frameworks because of its persistence and incomplete removal during treatment. Field studies in surface waters report an 18-hour half-life under combined photolytic and biotic conditions, although longer persistence estimates have also been reported (21, 27). Despite its environmental relevance, direct evidence on how desvenlafaxine interacts with bacterial competence and ARG acquisition remains limited. A closely related SNRI, duloxetine, has been shown to increase natural transformation of plasmid-borne ARGs in *Acinetobacter baylyi* ADP1 by up to several-fold at environmentally realistic concentrations (17). Because *A. baylyi* ADP1 is commonly found in wastewater and is phylogenetically very close to the clinically important pathogen *A. baumannii*, it is an established model for evaluating natural transformations (28, 29).

In this study, we therefore asked whether desvenlafaxine promotes natural transformation in A*. baylyi* ADP1 across concentrations that span and exceed those reported for WWTP effluents and hospital discharges. We first measured natural transformation using a plasmid as a proxy for environmental exDNA that carries antibiotic resistance markers to determine whether desvenlafaxine changes the frequency of ARG acquisition. We then tested envelope- and stress-related factors that could influence DNA uptake, including growth, membrane fluidity and permeability, ROS levels, surface charge, and quantified whether desvenlafaxine was cell-associated. Finally, we profiled global protein abundance changes by proteomics and used microscopy to examine competence-associated pili. Together, this workflow linked transformation outcomes to specific physiological and molecular changes that allowed us to propose a mechanism for desvenlafaxine’s effects.

## Methods

### Analysis of antidepressants in wastewater samples by LC-MS/MS

For this study, monthly water samples were collected from three WWTPs in New York, located near the Lake Erie-Niagara River corridor of the Laurentian Great Lakes Basin – representing urban (WWTP-1), urban and/or industrial (WWTP-2), and rural (WWTP-3) service areas – over a year (May 2024-May 2025). Sample preparation and cleanup were performed according to published methods (30, 31). Before sample collection, high-density polyethylene (HDPE) bottles were pre-rinsed with 10% (v/v) nitric acid. A sample volume of 500 mL was filtered through 1.7 μm pore-size GF/F Whatman glass microfiber filters, followed by 0.45 μm pore-size nylon membrane filters. The filtered samples are then acidified to pH 2.5 ± 0.5 using 85% phosphoric acid. Before the sample cleanup and pre-concentration stage, a 2 mL aliquot of Na_2_EDTA (5% w/v in water) was added to each sample and fortified with 50 μL of 1 mg/L mixture of 32 isotopically labeled surrogate standards. The fortified samples were subjected to solid-phase extraction (SPE) using tandem Hydrophilic-Lipophilic Balance (HLB) and mixed-mode cation exchange (MCX) cartridges, followed by liquid chromatography with tandem mass spectrometry (LC-MS/MS) analysis. Full details of the SPE procedure (including conditioning, elution solvents, evaporation, and reconstitution), chromatographic conditions (mobile phase composition and gradient program), mass spectrometry parameters, calibration curves, internal standard normalization, recovery corrections, and all quantification equations (including limits of detection and quantification) are provided in the supplementary Information (**Supplementary Method M1**).

### Bacterial strains, culture conditions and chemicals

A donor-recipient model of transformation was carried out to measure whether desvenlafaxine could promote the spread of ARGs via natural transformation. The naturally competent bacterium *A. baylyi* ADP1 (BD413, American Type Culture Collection, [ATCC] 33305, Manassas, VA, USA) was used as a recipient for the transformation experiment, as *Acinetobacter* spp. are frequently detected in the wastewater environment (32, 33). The shuttle plasmid pWH1266 (8.9 kbps, ATCC 77092) carrying two antibiotic-resistant gene cassettes (*tetA* against tetracycline and *blaTEM-1* against ampicillin) was selected as the donor. Plasmids were extracted from the host *E. coli* DH5α cells, grown at 37 °C in Luria-Bertani (LB) medium supplemented with 50 mg/L ampicillin, using Quick plasmid Miniprep Kit (Thermo Fisher Scientific, Waltham, MA, USA). The concentration of the extracted plasmid was measured by Nanodrop 2000 (Thermo Scientific, Waltham, USA). Tetracycline and desvenlafaxine (Catalog number: D2069-10MG) were purchased from Sigma-Aldrich (USA), while ampicillin was purchased from Gold Biotechnology (USA). Desvenlafaxine was dissolved in DMSO to reach 20mg/ml; further dilutions were carried out using MiliQ water.

### Natural transformation assay

The natural transformation assay was performed based on a previous study (17). The *A. baylyi* ADP1 cells were grown overnight at 30 °C in 5 mL LB broth with shaking at 200 rpm. Then, 50 μL of the overnight culture was transferred into 5 mL of fresh LB broth (1%, v/v) and further incubated for 6h at 30°C with shaking to reach the exponential growth phase (OD_600_ ∼1.1, ∼10^8^ cfu/mL). The cells were then collected and washed with 1X PBS (10X PBS: 137 mM NaCl, 2.7 mM KCl, 10 mM Na_2_HPO_4_, 1.8 mM KH_2_PO_4_, pH=7.2) twice using 6000·g for 5 min. Finally, the cells were resuspended in 1X PBS containing 50 mg/L sodium acetate (i.e., 36 mg/L chemical oxygen demand [COD] to mimic organics concentrations in untreated domestic wastewater (34). For each transformation assay, pWH1266 plasmid (A260/280 = 2.0; A260/230 = 1.8) was added into 500 μL of the resuspended *A. baylyi* ADP1 cells to a final plasmid concentration of 0.8 ng/μL. Then, desvenlafaxine was dosed into each of the 500 μL assays to obtain different final concentrations (control: 0 mg/L; low-level: 0.01, 0.1, and 1 mg/L; medium level: 10 mg/L; high-level: 50 mg/L) and incubated for 6h at 25 °C without shaking (n=6). The spot plating method was used, applying 20 μL spots on agar plates with at least five technical replicates, to count both transformants and total recipient cells. The transformant cells were counted on LB agar containing 100 μg/mL ampicillin and 5 μg/mL tetracycline after a maximum of 48 h incubation. In parallel, the total recipient cell number was counted on non-antibiotic-containing LB agar plates. Transformation frequency was measured by dividing the number of transformants by the total number of recipient cells as indicated in equation (1). Fold changes in transformation frequencies in desvenlafaxine-treated groups were calculated relative to untreated controls [equation (2)].

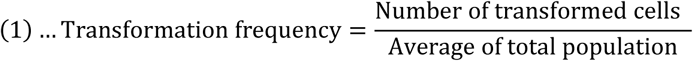

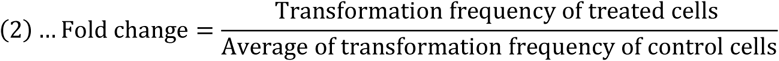

Transformants were confirmed by randomly selecting colonies from selective plates, followed by colony PCR and analysis via gel electrophoresis. Detailed information on the primers and thermocycling conditions is listed in **Supplementary Tables S1** and **S2**.

### Growth measurements and growth-rate analysis

Cultures of *A. baylyi* ADP1 were prepared by inoculating 5 mL of LB or minimal medium and incubating overnight (16 h) at 30 °C with shaking. The overnight culture was diluted 1/100 (500 µL culture into 4.5 mL fresh medium) to achieve an initial OD₆₀₀ ≈ 0.01. Desvenlafaxine was dissolved in Milli-Q water at a stock concentration of 10 mg/mL. Serial 2-fold dilutions were prepared in the same medium to generate 2× working concentrations ranging from 4000 mg/L down to 0.12 mg/L (final tested concentrations in the assay: 2000 mg/L to 0.06 mg/L). No DMSO was used as solvent. For each condition, 80 µL of the diluted bacterial suspension was aliquoted into triplicate wells of a 96-well plate, followed by 80 µL of the corresponding 2× drug concentration. Plates were incubated in a plate reader (Bioscreen C° Pro, Oy Growth Curves Ab Ltd, Helsinki, Finland) at 30 °C with continuous shaking. Optical density (OD₆₀₀) was measured every 15 min for 24 h. Specific growth rates (μ) were estimated from log-transformed, blank-corrected OD measurements using rolling-window linear regression. Growth rates were normalized to untreated controls, and growth inhibition or stimulation was quantified relative to the control. Because growth responses were non-monotonic and exhibited hormesis, growth-rate–based MICₓ values were defined on the inhibitory branch of the dose–response curve. Detailed descriptions of growth-rate estimation and MICₓ determination are provided in the **Supplementary Methods M2**.

### Measurement of ROS and cell membrane permeability

After exposure to desvenlafaxine, bacterial ROS generation and cell membrane permeability were measured and analyzed by flow cytometry (CytoFLEX Flow Cytometer, Beckman Coulter, USA). Cells were first prepared to reach an OD ∼1.1 in 1X PBS (∼10^8^ cfu/mL), similar to the transformation assay. Then the cells were diluted 100 times to reach ∼10^6^ cfu/mL. To determine ROS levels, cells were stained with 5uM of 2^/^,7^/^ -dichlorofuorescein diacetate (DCFDA) dye (Invitrogen, Thermo Fisher Scientific, USA) at 37 °C for 30 min. Cell aliquots (500 μL) were prepared in polystyrene flow tubes. Plasmid and desvenlafaxine were added to obtain different final concentrations (i.e., low, medium, and high) as mentioned previously, and incubated at 25 °C for 30 min. To determine cell membrane permeability, 2mM of propidium iodide (PI, Life Technologies, USA) was added to the same tubes by gentle pipetting, followed by incubation at 25 °C for 30 min in the dark. Fluorescence was detected by the flow cytometer. Fold change of ROS levels or cell membrane permeability was calculated by normalizing the desvenlafaxine-treated samples to those untreated controls. A 1.5% H_2_0_2_ was used as the positive control for ROS induction, while heating at 80 °C for 2h was used as the control for the confirmed membrane-permeable dead cells. All experiments were performed in at least three biological replicates.

### Cell Membrane fluidity measurement

Membrane fluidity of the treated *A. baylyi* cells was assessed by the polarization potential of a probe molecule (1,6-Diphenyl-1,3,4-hexatriene [DPH] dissolved in dimethylformamide [DMF], Sigma, St. Louis, USA) while embedded in the membrane and reacting to a polarized light in a fluorescent assay (35). Briefly, cells were prepared to reach an OD ∼1.1 in 1·PBS (∼10^8^ cfu/mL), like the transformation assay. Cells were then separated into tubes of DPH vs. no-DPH control. Plasmid and desvenlafaxine were added to each reaction, with desvenlafaxine tested at final concentrations of 10 mg/L and 50 mg/L. The reactions were kept for 6h at 25 °C without shaking in triplicate. DPH was then added to the DPH-labeled tubes at a final concentration of 0.4 µM in phosphate-buffered saline (1· PBS, pH 7.00±0.02) at a cell density of OD_600_ ∼0.5. Samples were then aliquoted into black-bottomed Nunclon Delta Surface 96-well plates. A plate reader (Synergy Multi-Mode microplate reader, BioTek) was used to measure polarization with a 360/40 nm (excitation) filter in a vertical position and a 460/40 nm (emission) filter in the vertical (I_VV_) or the horizontal (I_VH_) position for 15 minutes at 30 °C. Polarization was calculated using equation (3) (36), where G is the grating factor specific to the instrument, defined by the ratio of I_VV_ and I_VH_. This G-value was previously calculated for this instrument and was considered constant at 1.102. Within intact cell membranes, an increase in fluidity corresponds to a decrease in polarization values. Fold change analysis was done by normalizing the desvenlafaxine-treated samples to those untreated controls.

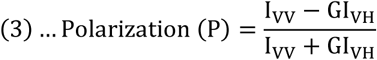

### Cell surface-charge measurement

Cell surface charge was measured by zeta potential as described previously (Soon et al., 2011). Briefly, bacterial cultures were grown to an optical density (OD) of approximately 1.1 in 1× PBS (∼10⁸ CFU/mL). Cells were washed twice and resuspended in Milli-Q™ water, followed by a 10-fold dilution in the same medium prior to transformation assays. Zeta potential was measured immediately after sample preparation (without the 6-hour incubation step) at final drug concentrations of 10 mg/L and 50 mg/L. The cell suspensions were loaded into disposable zeta cells (ATA Scientific, Australia) and analyzed using a Zetasizer Nano ZS™ instrument (Malvern Instruments Ltd., Malvern, UK) at 25 °C and 150 V. Measurements were conducted in triplicate, and electrodes were rinsed thoroughly with ethanol and Milli-Q™ water between each run.

### LC-MS/MS quantification of cell-associated drug levels

After transformation, 500 μL of bacterial suspension (OD ∼2) were centrifuged at 2,000 × *g* for 15 min, followed by removal of the supernatant and resuspension of the pellets in PBS. This wash cycle was repeated three times to remove extracellular drug and debris. Control media were processed similarly without cells. A 5 μL of internal standard (0.025 mg/mL in 1:8 methanol:water) was spiked into both cell pellets and cell-free control samples. Then, 400 μL of 8:2 methanol:water was added, followed by vortexing and incubation on ice for 10 min. Next, 800 μL mobile phase solution (5 mM of ammonium formate with 0.1% formic acid in nanopure water and 5 mM of ammonium formate with 0.1% formic acid in methanol) was added, tubes vortexed, sonicated for 10 min in water bath, vortexed again for 10 min, and centrifuged at max speed for 7 min at 4 °C. Supernatants were filtered (0.2 μm PTFE if needed) and transferred to glass LC-MS/MS vials. LC-MS/MS analysis and quantification of desvenlafaxine concentrations were performed using the same instrumental conditions, gradient elution profile, and calculation as described previously for the WWTP samples in the **Supplementary Method M1**.

### Protein extraction and proteomics

To study the translational responses upon desvenlafaxine exposure, another set of transformation assays with a larger volume (2 mL) was established at final concentrations of 10 mg/L and 50 mg/L. After 6hr, cells from each treatment group (n=3) were placed on ice for 15 min and harvested by centrifugation at 14,000 x g for 10 min at 4°C and washed 2x times with an ice-cold 1x PBS buffer pH 7.0. Cells were resuspended in 500 μL lysis buffer (1 Tablet of Complete Protease inhibitor [Roche #65726900] + 50mL 1X PBS buffer pH 7.0 + 1% sodium deoxycholate) and then sonicated for 5 minutes (pulse on for 10 seconds, pulse off for 20 seconds, amplitude of 75%) at 4°C. The lysates were centrifuged at 14,000 x g for 5 min at 4°C, and the supernatants were collected to determine the protein concentration using the Modified Lowry Protein Assay Kit (Thermo Scientific, USA). Total protein amounts were adjusted to 30 µg per sample. The lysates were denatured with Laemmli sample buffer containing 5% β-mercaptoethanol and heated for 8 min at 98°C. These treated samples were then run on SDS-polyacrylamide precast gels (Biorad #4561083) by electrophoresis at 120V. The gel was stained with Coomassie Brilliant Blue and de-stained according to standard protocols. Band intensities were qualitatively compared to visualize banding patterns in treatments vs. control (**Supplementary Figure S1**).

Total protein samples (70 ug proteins/sample) were then submitted to the proteomics facility of Iowa State University. Samples underwent reduction, alkylation, trypsin/Lys-C digestion, desalting, and spike-in with PRTC internal standard, followed by liquid chromatography–tandem mass spectrometry (LC-MS/MS) on a Thermo Scientific Orbitrap Astral mass spectrometer coupled to a Vanquish NEO HPLC system. Protein identification and label-free quantification (LFQ) were performed using Proteome Discoverer 3.2 with Mascot/Sequest HT (DDA mode) or CHIMERYS (DIA mode) search algorithms. PRTC-normalized peptide/protein abundances were exported from Proteome Discoverer 3.2, cleaned manually, and uploaded into MetaboAnalyst 5.0 for one-factor differential expression analysis. Data were processed with missing value imputation using k-nearest neighbors (KNN), initial filtering (sliders set to 0), and log 10 transformation normalization. Differentially expressed proteins were identified using an unpaired t-test with a raw significance threshold of p < 0.05. Full experimental protocols, instrument parameters, and raw data file details are provided in **Supplementary Methods M3** and the deposited mass spectrometry data to the ProteomeXchange Consortium via the PRIDE (37) partner repository with the dataset identifier PXD077476.

Proteins meeting the MetaboAnalyst significance criterion for each dose (10 mg/L vs control; 50 mg/L vs control) were used as input for protein–protein association analysis in STRING (STRING database, version 12.0). Significant proteins were queried using their UniProt accessions (or mapped from gene/locus tags when needed), with the organism set to *Acinetobacter baylyi* (strain ADP1) and default settings (medium confidence interaction, 0.4) for active interaction sources. Networks were visualized using STRING default confidence settings. STRING-predicted association clusters were then manually labeled based on the functional annotations.

### Electron microscopy study

Carbon film grids (Electron Microscopy Sciences, Hatfield, PA, USA) were prepared using a Pelco easiGlow Glow Discharge unit (Ted Pella, Inc., Redding, CA, USA) to create a hydrophilic surface. Transformed cells in 1X PBS buffer were applied to the grid (2 µL) and allowed to absorb for 30 seconds. Then, they were gently wicked with filter paper to form a thin film. Uranyl acetate stain (2% aqueous, 2 µL) was immediately applied to the film, allowed to sit for 30 seconds, and then gently wicked with filter paper before air-drying. Grids were evaluated and imaged using a JEOL JSM 2100 scanning transmission electron microscope (Jeol USA, Inc., Peabody, MA, USA) at 200kV with a Gatan OneView camera (Gatan. Inc., Pleasanton, CA, USA).

### Data analysis and statistics

All experiments were conducted independently in at least biological triplicate, unless otherwise mentioned. All data were expressed as mean ± standard deviation. Significant differences between the treated/control groups were analyzed using independent sample t-tests. P-values were corrected by the Benjamini-Hochberg correction method for multiple comparison. P_adj_ values less than 0.05 were considered to be statistically significant. All analyses and graphs were generated using R software (version 4.3.3). In select cases, Python (version 3.13) was used for specific visualizations, curve fitting, or supplementary calculations, as indicated in the relevant figure legends, text descriptions, or supplementary materials.

## Results

### Desvenlafaxine dominated antidepressant occurrence in WWTP effluents

Sampling was conducted at three municipal WWTPs in western New York (**Fig. 1A**). Among the antidepressants quantified from the effluents, a subset (i.e., desvenlafaxine, venlafaxine, bupropion, and sertraline) was detected in every sample (100% detection frequency across WWTPs; **Fig. 1B, Supplementary Table S3**). Concentration distributions differed by compound and by WWTP, but desvenlafaxine showed clear differences in magnitude and variability across sites (Fig. 1B). At the first plant, concentrations ranged from 460 ppt to 0.7 ppm, with a median of 3,800 ppt., influenced by two outliers that substantially extended the upper limit. The second plant had comparatively lower concentrations (158–4,400 ppt), with a median of 1,400 ppt. Conversely, the third plant demonstrated higher overall concentrations (486–16,900 ppt), with the highest median of 9,100 ppt. Together, these findings confirm desvenlafaxine as the dominant antidepressant in WWTP effluent across the sampled region.

**Figure 1.**
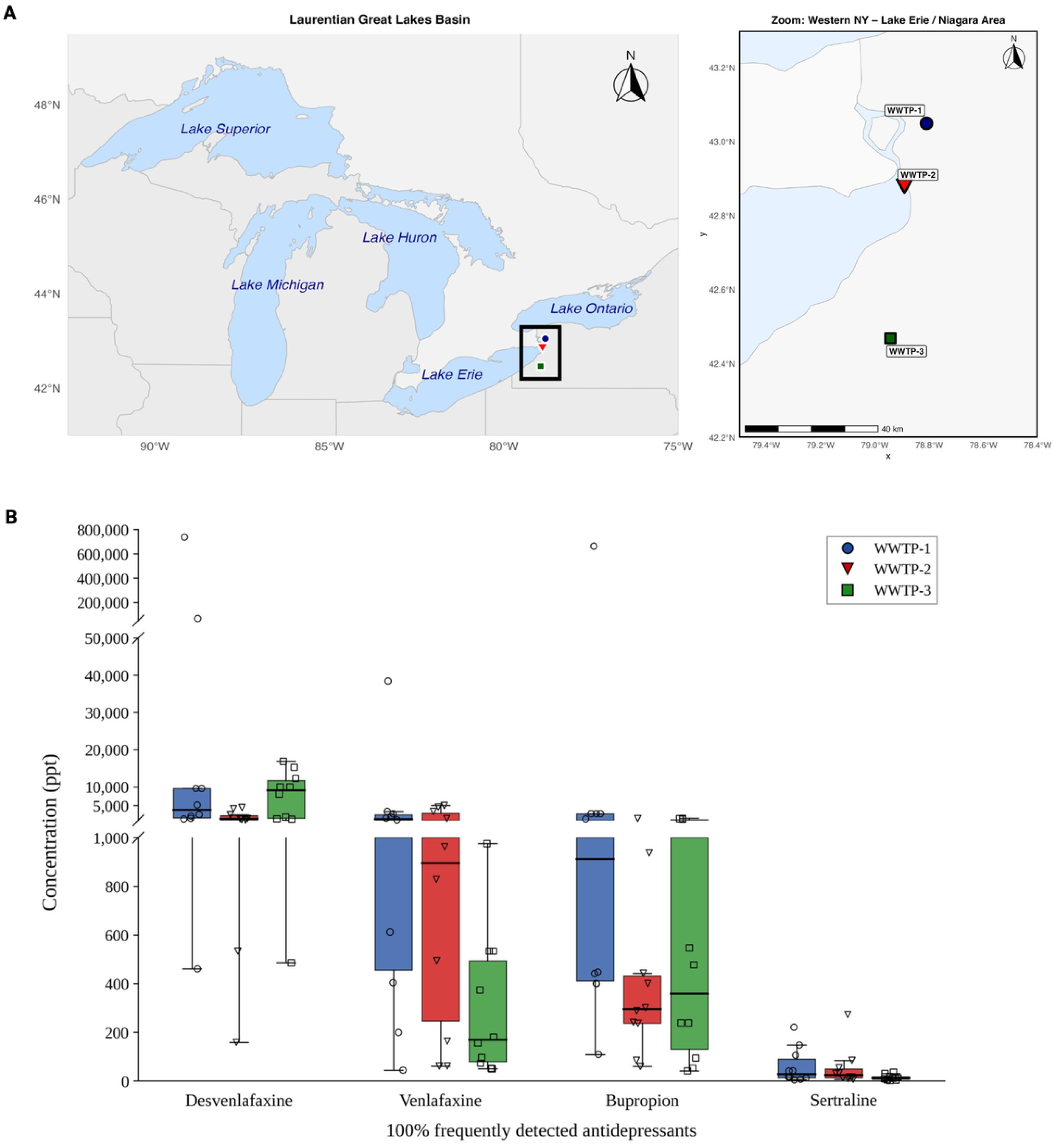
Desvenlafaxine was the most abundant of four antidepressants consistently detected in WWTP effluents from the Lake Erie–Niagara corridor. (A) Three municipal WWTPs were sampled in western New York: WWTP-1 (urban), WWTP-2 (urban and/or industrial), and WWTP-3 (rural). The left panel shows their location within the Laurentian Great Lakes Basin; the right panel provides a zoomed view. (B) Effluent concentrations (ppt) of the four antidepressants detected in 100% of samples. Boxplots show site-specific distributions; individual points represent monthly sampling events. The y-axis is broken to accommodate the wide concentration range.

### Desvenlafaxine induced transformation at mid to high doses tested

To assess whether desvenlafaxine promotes HGT, we used pWH1266 as a proxy for environmental ARG-bearing exDNA. Desvenlafaxine significantly increased the frequency of natural transformation in *Acinetobacter baylyi* ADP1, but only at certain doses (**Fig. 2A**). Across the tested range (0.01–50 mg/L), transformation frequency (fold change relative to the untreated control) remained close to baseline at the lowest concentrations (0.01–1 mg/L). The clear increases appeared at mid-range doses: ∼2.0-fold (i.e., 1.74 ± 0.33) at 10 mg/L and ∼1.5-fold (i.e., 1.49 ± 0.19) at 50 mg/L (P_adj_ < 0.05, Bonferroni correction, independent sample t-test vs. control). The effect was consistent across replicates.

**Figure 2.**
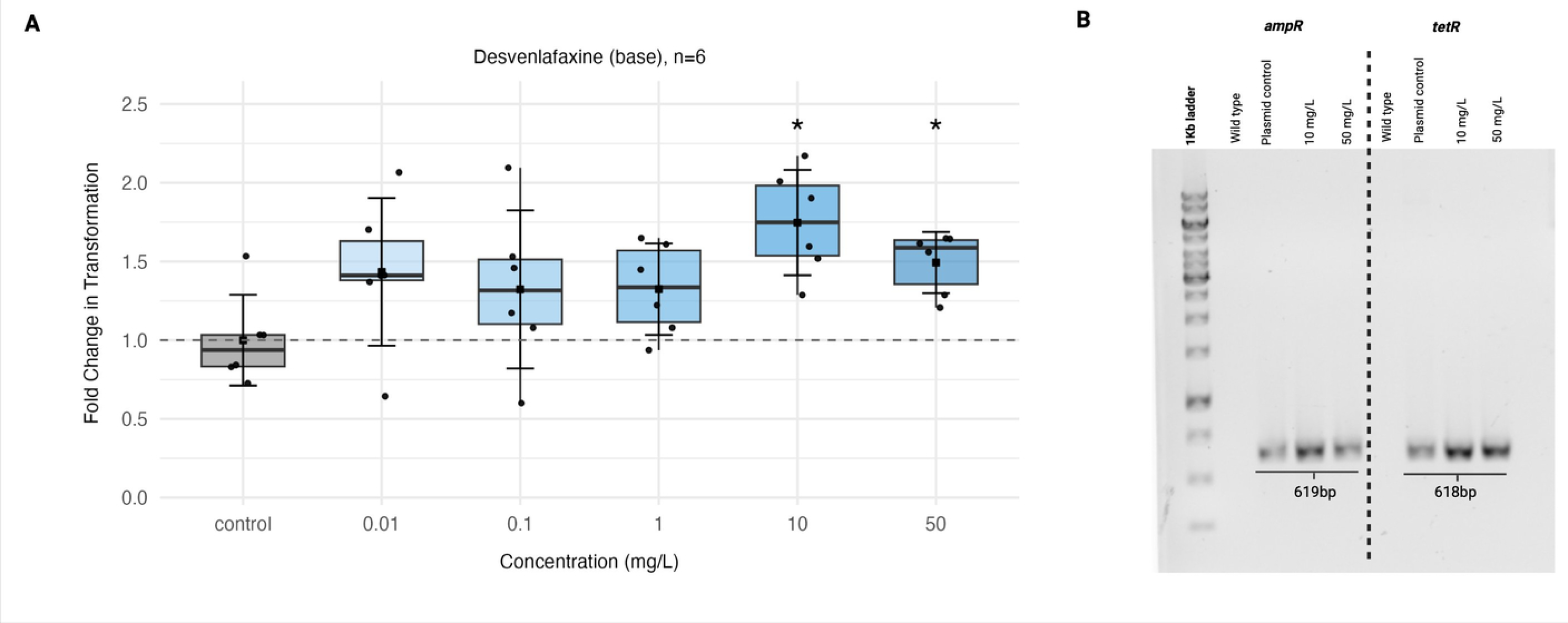
Desvenlafaxine increases natural transformation in *Acinetobacter baylyi* at medium-to-higher doses tested. (A) Fold change in transformation frequency relative to the untreated control after exposure to desvenlafaxine (0–50 mg/L). Boxplots show the median and interquartile range, with individual points representing biological replicates (n = 6 per condition). The dashed line indicates the baseline (fold change = 1, no change relative to control). Asterisks indicate significant differences relative to the untreated control (*P∼adj∼ < 0.05, Bonferroni-corrected independent t-tests). (B) PCR confirmation of transformants carrying plasmid markers. Representative gel showing amplification of ampR (619 bp) and tetR (618 bp) in transformant colonies at 10 mg/L and 50 mg/L desvenlafaxine compared with wild-type and plasmid-positive controls; 1 kb ladder shown at left.

To confirm that the observed transformants resulted from true plasmid uptake rather than spontaneous mutation or contamination, we ran PCR on the transformants using primers for the ampicillin (*ampR*, 619 bp) and tetracycline (*tetR*, 618 bp) resistance cassettes. Gel electrophoresis showed bands at the expected sizes for both cassettes in transformants recovered from the 10 mg/L and 50 mg/L exposures (**Fig. 2B**). The plasmid control yielded the same bands, while wild-type *A. baylyi* (no plasmid) showed no amplicons, confirming the absence of endogenous resistance genes and validating the assay’s specificity.

### Desvenlafaxine was not toxic to *A baylyi* cells

We next examined whether desvenlafaxine caused any toxicity or stress that could explain the transformation effect, so we tested growth kinetics (0.06-2000 mg/L) (**Fig. 3**), membrane permeability, and ROS levels. Using the maximum specific growth rate (μ_max_) normalized to the untreated control (μ_0_), growth rates remained close to control values from 0.06 to 500 mg/L (μ_max_/μ_0_ approximately 0.93–1.00 across most concentrations) (**Fig. 3A**). Growth inhibition became evident only at the highest concentrations tested, with μ_max_/μ_0_ decreasing to ∼0.83 at 1000 mg/L and ∼0.67 at 2000 mg/L (corresponding to ∼17% and ∼33% inhibition, respectively). Dose–response analysis of growth inhibition yielded an estimated concentration for 20% inhibition (i.e., IC20) of approximately 1353 mg/L (**Fig. 3B**). Together, these results indicate that desvenlafaxine does not cause substantial growth toxicity to *A. baylyi* under our conditions at low-to-moderate concentrations, and that measurable inhibition occurs only at very high concentrations.

**Figure 3.**
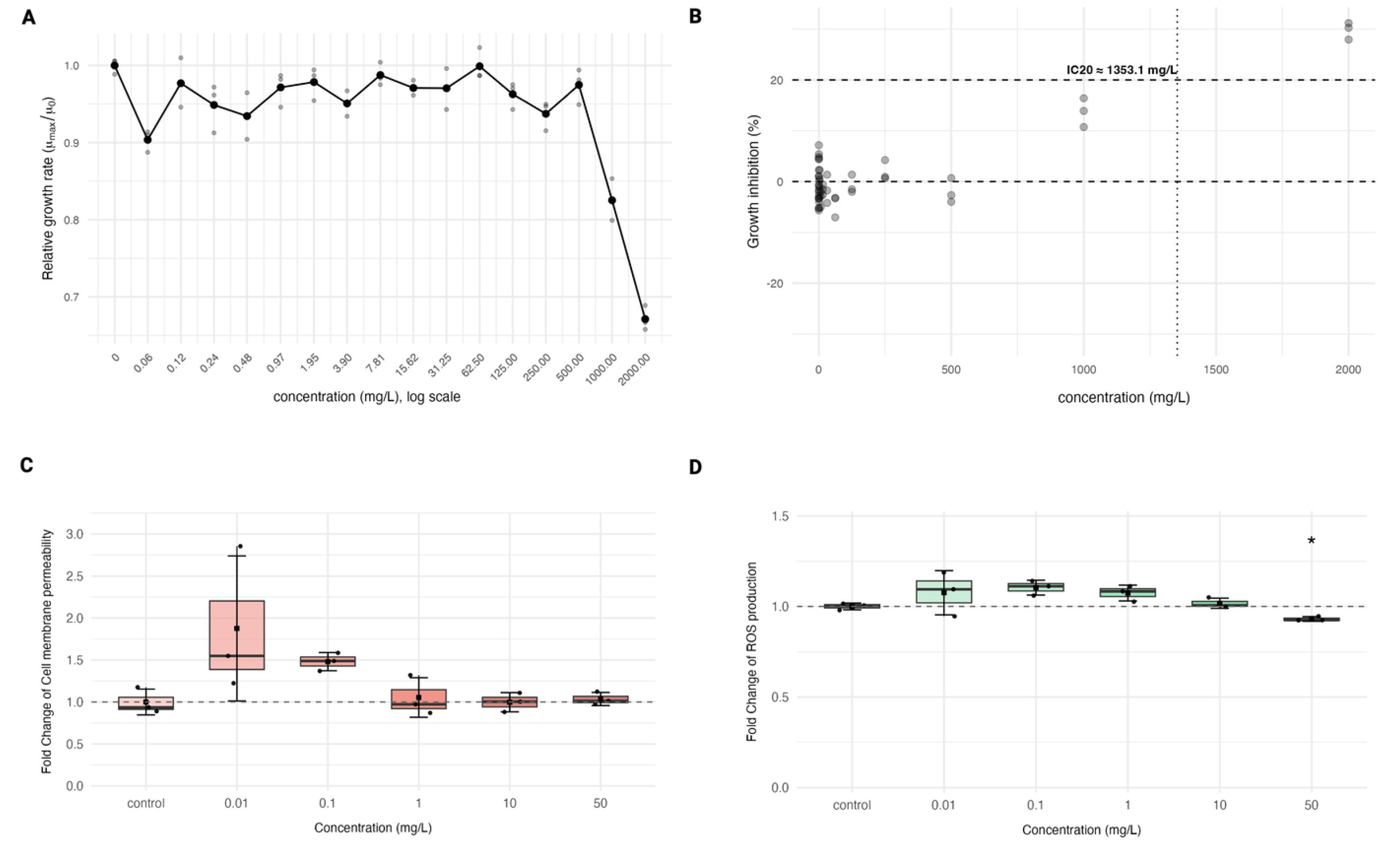
Minimal toxicity to *A. baylyi* cells observed during desvenlafaxine treatment. (A) Maximum specific growth rate (μmax) calculated from growth curves and normalized to the untreated control (μ0), plotted versus desvenlafaxine concentration (log scale). Points represent mean and gray symbols indicate individual biological replicates across independent experiments. (B) Growth inhibition (%) derived from normalized growth-rate measurements; dashed line indicates 20% inhibition and vertical marker indicates the estimated 20% inhibitory concentration (“IC20” ≈ 1353.1 mg/L). (C) Cell membrane permeability measured as fold change relative to the untreated control after exposure to desvenlafaxine (0–50 mg/L). (D) Intracellular ROS production measured as fold change relative to the untreated control across the same concentration range. Boxplots show the median and interquartile range, with individual points representing biological replicates (**n = 3** per condition). Statistical comparisons were performed using pairwise t-tests with Bonferroni correction; ns indicates no significant difference relative to the control. The dashed line indicates the baseline (fold change = 1, no change relative to control). Asterisks indicate significant differences relative to the untreated control (*P∼adj∼ < 0.05, Bonferroni-corrected independent t-tests).

Flow cytometry was used to assess whether desvenlafaxine exposure altered membrane permeability or induced ROS production in *A. baylyi*. Membrane permeability, measured by propidium iodide uptake, showed no significant change across the tested concentrations (**Fig. 3C**). At all doses from 0.01 to 50 mg/L, the distribution of fluorescence intensity remained similar to the untreated control, with median values near baseline and no detectable shift toward higher permeability. Similarly, ROS levels, monitored using the DCFH-DA probe, displayed no evidence of induction by desvenlafaxine (**Fig. 3D**). When comparing the untreated control to exposed cells, significance in median fluorescence signal was not detected at each of the concentrations (0.01, 0.1, 1, 10, and 50 mg/L). These results indicate that desvenlafaxine exposure did not lead to measurable increases in membrane permeability or ROS generation under the conditions tested.

### Cell-associated desvenlafaxine increases membrane fluidity and reduces surface negativity

Since desvenlafaxine did not induce ROS or membrane permeability changes, we next investigated whether it instead affected other membrane properties, specifically, membrane fluidity and surface charge that could facilitate DNA uptake. When cells were exposed to desvenlafaxine, we observed clear, dose-dependent shifts in their membrane properties and surface electrostatics (**Fig. 4**). For instance, membrane fluidity increased markedly, as shown by a significant drop in DPH fluorescence anisotropy from 0.457 ± 0.02 in untreated controls to 0.238 ± 0.01 at 10 mg/L and further to 0.162 ± 0.03 at 50 mg/L (n = 3 per group; **Fig. 4A**, ***p_adj_ <0.001 vs. control) (lower anisotropy means higher fluidity). At the same time, the cells’ surface became less negatively charged, with zeta potential values moving from −23.77 ± 0.38 mV in controls to −21.80 ± 0.56 mV at 50 mg/L (n = 3 per group; **Fig. 4B**; ** p_adj_ <0.01 vs. control). The changes were not significantly different at 10 mg/L.

**Figure 4.**
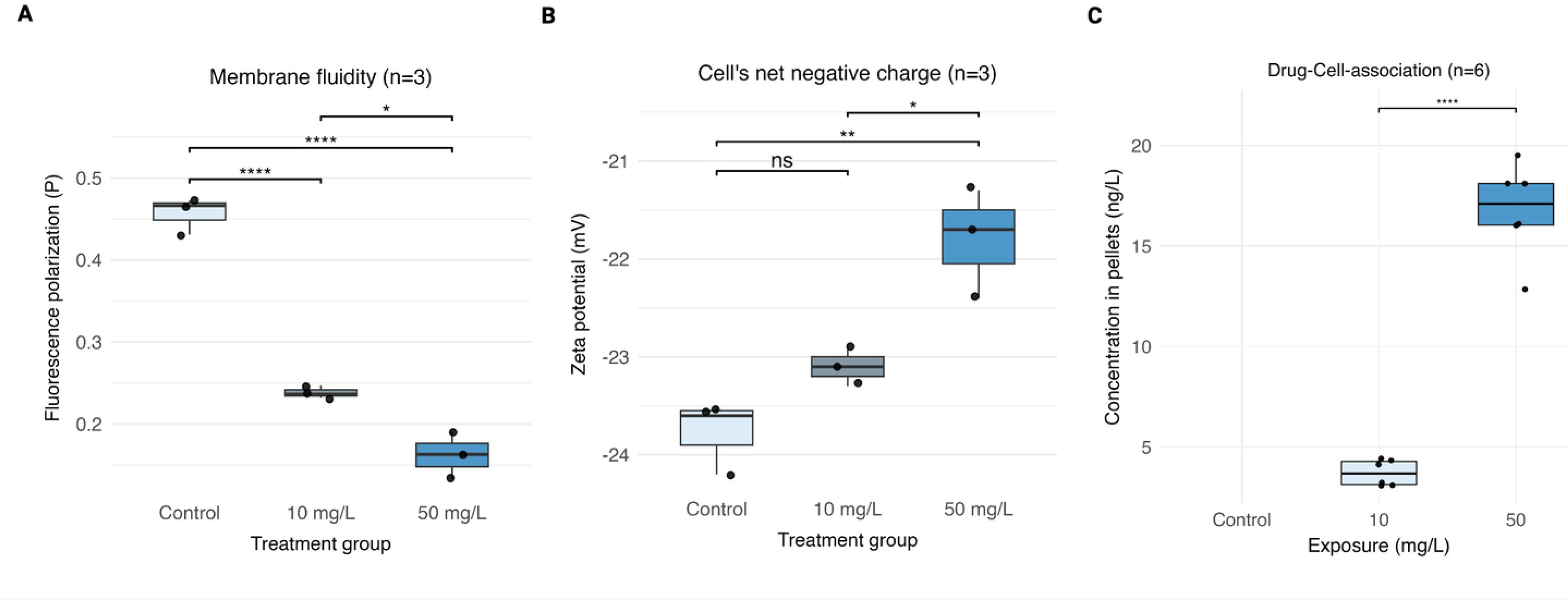
Desvenlafaxine increases membrane fluidity, reduces net negative surface charge, and shows cell-associated accumulation. (A) Membrane fluidity measured by DPH fluorescence anisotropy following exposure to desvenlafaxine (0, 10, and 50 mg/L). Lower anisotropy indicates higher membrane fluidity. (B) Cell surface charge measured as zeta potential (mV) after the same exposures. Less negative values indicate reduced net negative surface charge. (C) Cell-associated desvenlafaxine quantified in the cell fraction (ng/L) after exposure to 10 and 50 mg/L. Boxplots show the median (center line) and interquartile range (box), with whiskers indicating the data range; individual points represent biological replicates. Statistical significance in (A–B) was assessed using pairwise t-tests with Bonferroni correction; significance in (C) was assessed by an unpaired two-sample t-test. ns, not significant; * p_adj < 0.05; ** p_adj < 0.01; *** p_adj < 0.001; **** p_adj < 0.0001. Replicate numbers: (A–B) n = 3 per condition; (C) n = 6 per condition.

To confirm that these membrane changes could be directly attributed to the physical presence of desvenlafaxine, we also quantified drug’s localization using LC-MS/MS in washed pellets (**Fig. 4C**). Desvenlafaxine was mostly cell-associated, building up from 3.71 ± 0.65 ng/L at 10 mg/L to 16.78 ± 2.4 ng/L at 50 mg/L—about 4.5 times higher at the elevated dose (n = 6 per group; **Fig. 4C**; **** p < 0.0001 vs. control), corresponding to estimated intracellular concentrations of 15–60 nM. No drug signal was detected in no-cell controls (drugs in buffer only), which confirms our washing steps removed any extracellular drug below detectable levels (LOD of 4.7 ng/L) and that the observed membrane effects were driven by the drug interacting with the cells.

### Desvenlafaxine increased competence-related protein abundance at low doses

Proteomic analysis was performed on cells exposed to 10 mg/L or 50 mg/L desvenlafaxine compared to untreated controls (0 mg/L). Volcano plots revealed differential protein abundance patterns at each dose (**Figure 5A–B**). At 10 mg/L, 33 proteins were significantly increased (up) and 3 were decreased (down) based on fold-change and significance thresholds (**Fig. 5A**). In contrast, at 50 mg/L, 13 proteins were upregulated, and 4 were downregulated (**Fig. 5B**).

**Figure 5.**
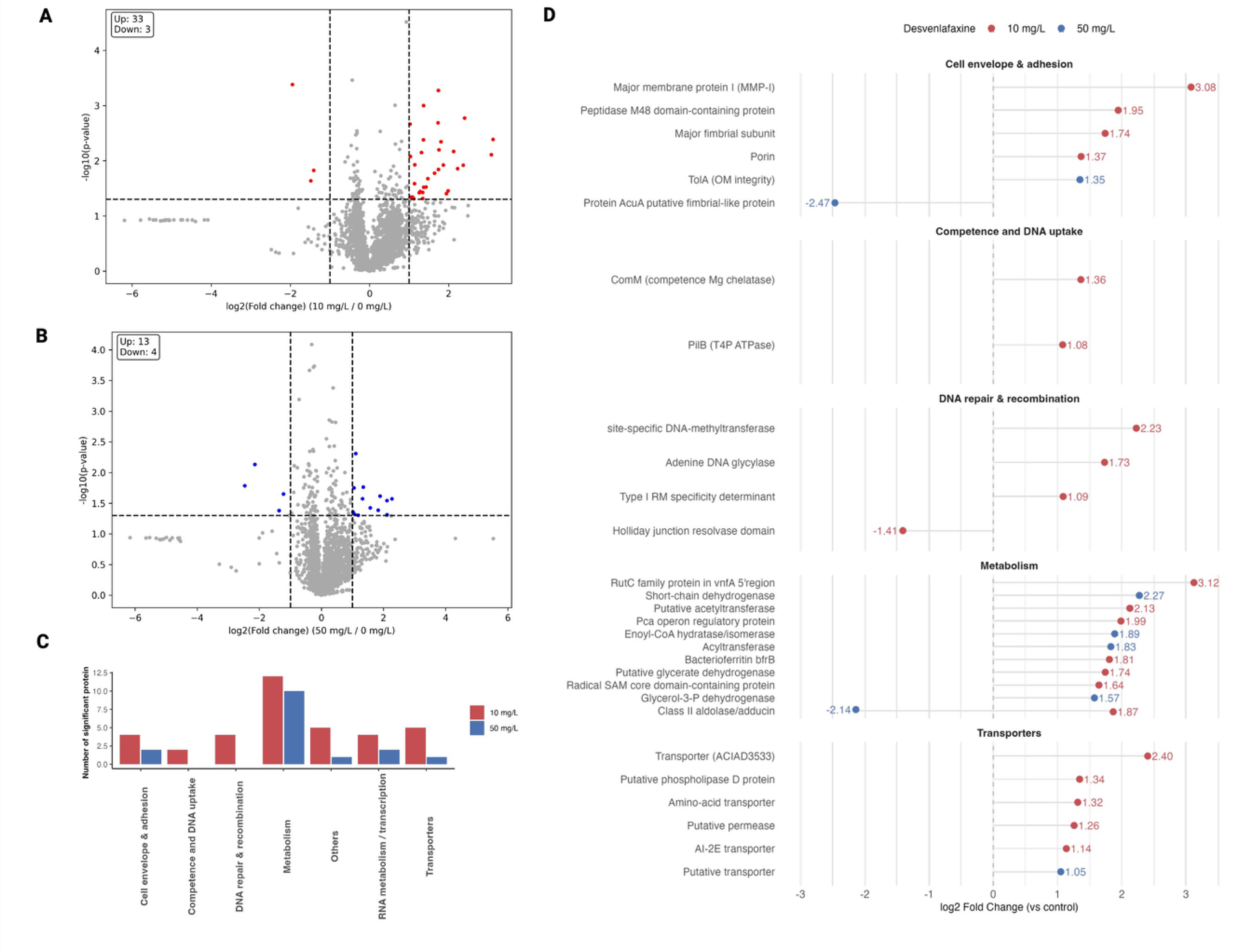
Desvenlafaxine competence-related protein abundance at low doses. (A–B) Volcano plots showing differential protein abundance in cells exposed to 10 mg/L (A) or 50 mg/L (B) desvenlafaxine relative to the untreated control (0 mg/L). The x-axis shows log2(fold change) and the y-axis shows −log10(p-value); dashed lines indicate the fold-change and significance thresholds used to define differentially abundant proteins. Points highlighted in color represent proteins meeting both thresholds, and the numbers in the inset indicate counts of proteins increased (“Up”) or decreased (“Down”) under each condition. (C) Functional distribution of significantly changed proteins, grouped into categories including cell envelope/adhesion, competence and DNA uptake, DNA repair/recombination, metabolism, RNA metabolism/transcription, transporters, and others, for 10 mg/L and 50 mg/L treatments. (D) Selected differentially abundant proteins plotted by log2 fold change (vs control) and organized by functional category, highlighting envelope-associated factors (e.g., porins/outer membrane integrity), competence/DNA uptake proteins, DNA repair/recombination proteins, metabolic enzymes, and transporters. Red points indicate 10 mg/L and blue points indicate 50 mg/L.

The significantly perturbed proteins were further categorized by functions (**Fig. 5C**). The largest number of changes occurred in metabolism under both doses. Proteins annotated under competence/DNA uptake and DNA repair/recombination were only observed at 10 mg/L. Within the competence and DNA uptake group at 10 mg/L, two proteins increased in abundance: ComM (competence Mg chelatase) (log2FC +1.36) and PilB (T4P ATPase) (log2FC +1.08) (**Fig. 5D**). Several proteins related to DNA repair/recombination also upregulated at 10 mg/L (e.g., site-specific DNA methyltransferase (log2FC +2.23)), alongside decreased abundance of a Holliday junction resolvase domain protein (log2FC −1.41) (**Fig. 5D**). Cell envelope and adhesion proteins were all upregulated at 10 mg/L, which included major membrane protein I (MMP-I) (log2FC +1.95), a peptidase M48 domain-containing protein (log2FC +1.74), and a porin (log2FC +1.37). In contrast, the 50 mg/L condition showed fewer envelope-associated changes among the selected proteins, with TolA (outer membrane integrity) increased (log2FC +1.35) and a putative fimbrial-like protein (AcuA) decreased (log2FC −2.47). Just one transporter was induced at 50 mg/L compared to five at 10 mg/L, led by the transporter ACIAD3533 (log2FC +2.40), followed by a permease and an AI-2E transporter. Among metabolic proteins, several enzymes involved in aromatic compound sensing were upregulated. The PCA operon regulatory protein (PcaU) was upregulated (log2FC, +1.99) at 10 mg/L, as was benA (benzoate 1,2-dioxygenase, not shown in the figure) (see full protein list in the **Supplementary Table S4**).

To determine whether the significant proteins were functionally related, we mapped them to the STRING database, which integrates known and predicted protein–protein interactions to identify shared biological processes (**Supplementary Fig. S2**). The proteins at 50 mg/L could not find any interactions based on the current STRING database. However, at the 10 mg/L drug concentration, multiple protein–protein interactions were identified. These proteins were manually grouped into five functional clusters based on their annotated biological roles (Table 1): (A) Membrane transport, (B) Iron homeostasis/Stress response, (C) Transcriptional regulation, (D) Envelope remodeling, and (E) Restriction-modification/DNA methylation. Complete STRING annotations for all proteins are provided in **Supplementary Table S5**.

**Table 1:** Proteins and STRING association clusters identified at 10 mg/L desvenlafaxine (functional network labels assigned manually).

| Association | Proteins involved |  | Abundance Increased (↑) or Decreased (↓) (Fig. 5D) | Functional cluster label* |
| --- | --- | --- | --- | --- |
|  | UniProt No. | Name |  |  |
| A | Q6F720 | Porin | ↑ | Membrane transport |
|  | Q6F7K3 | Holliday Junction resolvase | ↓ |  |
|  | Q6F7K1 | AI-2E family transporter | ↑ |  |
| B | Q6F7G7 | RutC family protein | ↑ | Iron homeostasis/Stress response |
|  | Q6FDV6 (bfrA) | Bacterioferritin A | ↑ |  |
|  | Q6F7G5 (bfrB) | Bacterioferritin A | ↑ |  |
| C | Q6FB58 | AraC family regulator | ↑ | Transcriptional Regulation |
|  | Q6F6W7 | TetR family regulator | ↑ |  |
| D | Q6FFT2 | dTDP-glucose-4,6-dehydratase | ↑ | Envelope remodeling (LPS/O-antigen biosynthesis) |
|  | Q6FFT7 | Glycyl transferase family 1 | ↑ |  |
| E | Q6F777 | Adenine-specific DNA methyltransferase | ↑ | Restriction-modification/DNA methylation |
|  | Q6F778 | Type I restriction-modification system | ↑ |  |

\*Clusters were identified using STRING v12.0 (medium confidence 0.4). Full raw STRING annotations are available in the deposited dataset.

Following the proteomic analysis, we used TEM to identify any changes in surface structures resembling competence-associated T4 pili. In representative images, thin filamentous structures (∼7 nm wide; arrows) were clearly visible on cells exposed to desvenlafaxine, but were only occasionally observed in untreated controls (**Fig. 6**). To summarize this pattern across images, pili frequency was scored semi-quantitatively across multiple magnifications (8K–25K) and pooled within each condition (**Table 2**). In the control group (33 cells examined), pili were generally absent—most fields showed none or only 1 or 2 sporadic pili (rated −/+). In contrast, both desvenlafaxine treatments showed noticeably more consistent piliation: at 10 mg/L (36 cells) and 50 mg/L (39 cells), cells averaged at least three pili per field (rated ++ for each). The scores lined up well across all magnifications, so the difference wasn’t driven by any single imaging scale. Together, these TEM images provide structural evidence that desvenlafaxine exposure was associated with increased detection of thin pili-like appendages relative to the untreated condition.

**Figure 6.**
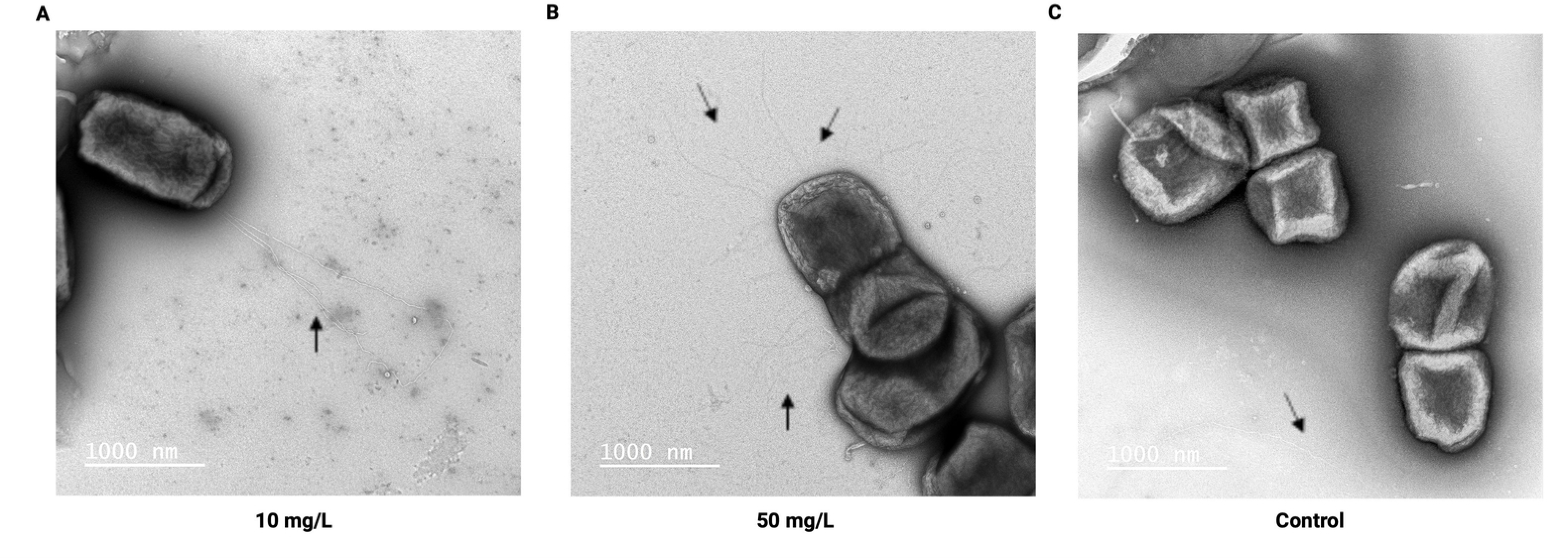
Desvenlafaxine exposure increases pili-like surface appendages in *Acinetobacter baylyi* ADP1. Transmission electron microscopy (TEM) images of negatively stained cells after 6 h exposure to desvenlafaxine or untreated control. (A) At 10 mg/L, thin filamentous structures (∼7 nm wide; arrow) consistent with type IV pili were consistently observed extending from the cell surface (polar distribution). (B) At 50 mg/L, pili-like appendages remained abundant and were distributed across the cell surface in a non-polar pattern (arrows). (C) In untreated control cells, pili were only sporadically detected (arrow). Scale bars = 1,000 nm. Semi-quantitative scoring across multiple magnifications (8K–25K) confirmed that both desvenlafaxine-treated conditions (10 and 50 mg/L) showed ≥3 pili per cell on average (rated ++), compared to absent or single sporadic detection in controls (rated −/+; see Table 2).

**Table 2:** Semi-quantitative evaluation of TEM negative staining of *A. baylyi* obtained under different desvenlafaxine conditions.

| Condition | Total cells evaluated | Relative frequency of 7-nm pili | Rating* (pooled) |
| --- | --- | --- | --- |
| Control | 33 | No pili or single/sporadic detection (except one field at low magnification) | – / + |
| 10 mg/L | 36 | $\geq 3$ pili per cell on average (consistent across all fields) | ++ |
| 50 mg/L | 39 | $\geq 3$ pili per cell on average (consistent across all fields) | ++ |

\*Ratings are defined as follows: – indicates no pili or only single detection; + indicates sporadic pili (or up to approximately 1 pilus per 25–50 cells); ++ indicates $\geq 3$ pili per cell on average. Scoring of pili frequency was performed independently at multiple magnifications (ranging from 8K to 25K) to minimize potential visibility bias arising from differences in resolution and field coverage; results were highly similar across all magnifications. Frequent clustering of bacteria on TEM sample grids prevented unbiased quantitative estimation of the distribution of pili per individual cell in some fields. Total cells evaluated represent the sum of bacteria counted across all images and conditions in the respective treatment group.

## Discussion

The potential role of antidepressants in promoting antibiotic resistance has received far less attention. This work provides the first evidence that desvenlafaxine, one of the most abundant antidepressants that occur at high frequency in municipal wastewater, can promote HGT by natural transformation in bacteria. It was consistently found at higher quantities than other antidepressants during our monitoring at three WWTPs in western New York (median effluent levels up to 9,100 ng/L at the WWTP-3; **Fig. 1B**). The result is consistent with a U.S. national survey, which found desvenlafaxine in 35% of freshwater ecosystems, with maximum dissolved concentrations of about 4600 ng/L (38). These high environmental levels reflect both heavy usage and the poor removal of the drug by conventional treatment, much like its parent compound venlafaxine, which typically shows only 0–30% removal (39).

In this work, we found that desvenlafaxine enhances the transformation of a multidrug resistance plasmid into *A. baylyi* ADP1. The effect was modest but reproducible, with a ∼2-fold increase at 10 mg/L and ∼1.5-fold at 50 mg/L. Although transformations at lower concentrations (≤ 1mg/L) were not statistically significant, these concentrations can produce small shifts that are difficult to resolve with biological variability, yet may become more consequential during chronic exposure or in the presence of co-stressors (e.g., other pharmaceuticals, metals, disinfectants etc.) typical of wastewater environments (13). Importantly, the highest desvenlafaxine level we measured in one of the WWTPs reached 0.7 mg/L. This suggests that environmentally relevant exposures could plausibly contribute to incremental increases in plasmid uptake potential in situ. Our findings extend prior work on non-antibiotic drivers of HGT. For example, Lu *et al.* (2022) showed that several antidepressants (e.g. sertraline, duloxetine, fluoxetine, bupropion) can promote *A. baylyi* transformation (17). Likewise, Ding *et al.* (2022) reported that SSRIs and SNRIs accelerate conjugative transfer of multidrug plasmids across species (40). In both cases, enhanced HGT was linked to oxidative stress, altered membrane permeability, and upregulation of general stress pathways. In contrast, desvenlafaxine’s effect appears to function through a distinct route. Under our growth conditions, it was generally non-toxic (IC20 = 1353 mg/L; **Fig. 3B**) and did not raise ROS or conventional permeability (no PI uptake shift; **Fig. 3**). Thus, unlike other antidepressants, desvenlafaxine enhanced transformation without obvious cell damage or stress.

Our biophysical measurements suggest that the positively charged amine group of desvenlafaxine interacts directly with the bacterial envelope. Upon exposure, *A. baylyi* membranes became significantly more fluid (lower DPH anisotropy; **Fig. 4A**) and the cell surface charge became less negative (zeta potential shifted from –23.8 to –21.8 mV at 50 mg/L; **Fig. 4B**). These changes imply that desvenlafaxine partitions into the lipid bilayer and neutralizes anionic surface groups. This is analogous to the well-known action of cationic antimicrobials: for example, polymyxin B (a cyclic cationic peptide) binds electrostatically to the negatively charged lipopolysaccharide (LPS) of Gram-negative cells, which neutralizes surface charge and destabilizes the outer membrane (41). Such interaction destabilizes the outer membrane, increasing permeability and altering membrane architecture. By analogy, desvenlafaxine’s cationic group likely interacts with LPS charges and reduces electrostatic repulsion to aid DNA binding. Indeed, prior studies have shown that any neutralization of surface charge (less negative zeta potential) often correlates with increased membrane permeability (41, 42). In our case, although the dye uptake did not change appreciably, even subtle membrane perturbations may permit greater plasmid contact or uptake. Increased membrane fluidity itself can also promote fusion and vesicle formation. Together, the net effect is that the drug-treated cells have more fusogenic, less repulsive surfaces. This would favor the electrophoretic attachment of negatively charged DNA and its uptake via the competence machinery. Notably, several antidepressants have been shown to disrupt membranes. Sertraline and paroxetine cause morphological changes and enhanced permeability in *E. coli* and *S. aureus*, as well as ROS production (43). Desvenlafaxine thus seems to clearly modulate membrane lipids to aid transformation.

Our proteomic and microscopic analyses support a model of enhanced competence. At 10 mg/L desvenlafaxine, *A. baylyi* upregulated multiple proteins involved in competence and DNA uptake (**Fig. 5D**). The competence-associated helicase ComM and the type IV pilus extension ATPase PilB were both more abundant. ComM has been shown to form a hexameric helicase that promotes branch migration of transforming DNA during uptake (44), so its induction suggests more active DNA recombination. PilB is a key extension ATPase motor of type IV pili (T4P) systems in *A. baylyi* (44). Thus, upregulation of PilB is consistent with increased pilus formation. Accordingly, TEM of negatively stained cells revealed many thin, pili-like filaments (∼7 nm) on drug-treated cells than on controls (**Fig. 6**, Table 2). However, at 50 mg/L, these proteins were not detected, even though TEM still indicated abundant pili-like filaments. This suggests that pili abundance alone may not result in maximal DNA uptake, because T4P must retract to pull bound DNA to the cell surface (45). In *A. baylyi*, pilus function depends on a balance of motor activities that control both extension and retraction; higher-dose exposure may shift cells toward a general stress state that favors pilus retention rather than an optimized competence (44). Together, these data indicate that desvenlafaxine stimulates the structural components of competence (more pili) and, at moderate exposure, also the enzymatic machinery (ComM) needed for efficient DNA internalization and integration.

Interestingly, several proteins tied to aromatic metabolism (i.e., PcaU and BenA) were upregulated at 10 mg/L. These proteins feed into the β-ketoadipate pathway (46, 47). PcaU is a protocatechuate-dependent activator of the pca-qui operon, and the ben operon (*benABCDE*, including BenA) initiates benzoate catabolism. Their co-induction implies that *A. baylyi* senses desvenlafaxine as an aromatic-like substrate (xenobiotics), turning on catabolic genes (48). By contrast, no aromatic-regulator proteins were induced at 50 mg/L, again indicating that low-dose drug specifically triggers an aromatic nutrient response. Also, at 10 mg/L, we observed more transporters significantly expressed (five transporters upregulated vs. one at 50 mg/L; **Fig. 5C**). In parallel, LC–MS detected higher cell-associated accumulation of the drug at 50 mg/L than at 10 mg/L (**Fig. 4C**). Since this assay, according to our protocol, can’t distinguish cytosolic vs. surface drug, this may suggest that 10 mg/L had better uptake/efflux regulation, while 50 mg/L caused excessive accumulation that overwhelms these responses. It is consistent with the general principle that bacterial intracellular small-molecule levels reflect a concentration-dependent permeability–efflux balance (49). Alternatively, these proteins may have been transiently expressed at 50 mg/L but already returned to baseline by the 6-hour sampling point, reflecting accelerated kinetics rather than suppression. Future studies across multiple time points would help distinguish these possibilities. The STRING network analysis further grouped 10 mg/L-induced proteins into clusters for membrane transport, envelope remodeling (LPS synthesis enzymes), iron-homeostasis/stress response, and transcriptional regulation (Table 1). For example, upregulation of a porin together with an AI-2E family transporter likely does not substitute for the PilB-driven competence machinery. Rather, it suggests broader changes in membrane transport. Increased porin abundance can raise permeability to small solutes/ions, which may shift ionic balance and membrane potential and contribute to envelope remodeling (e.g., altered membrane fluidity) (50). Alternatively, at 50 mg/L, no network was identified. This biphasic proteomic response implies that moderate doses (i.e., 10 mg/L) trigger a coordinated metabolic adaptation, whereas higher doses (i.e., 50 mg/L) may partially shut down or overwhelm these pathways. Overall, **Fig. 7** proposes the putative mechanisms by which antidepressants promote natural transformation.

**Figure 7.**
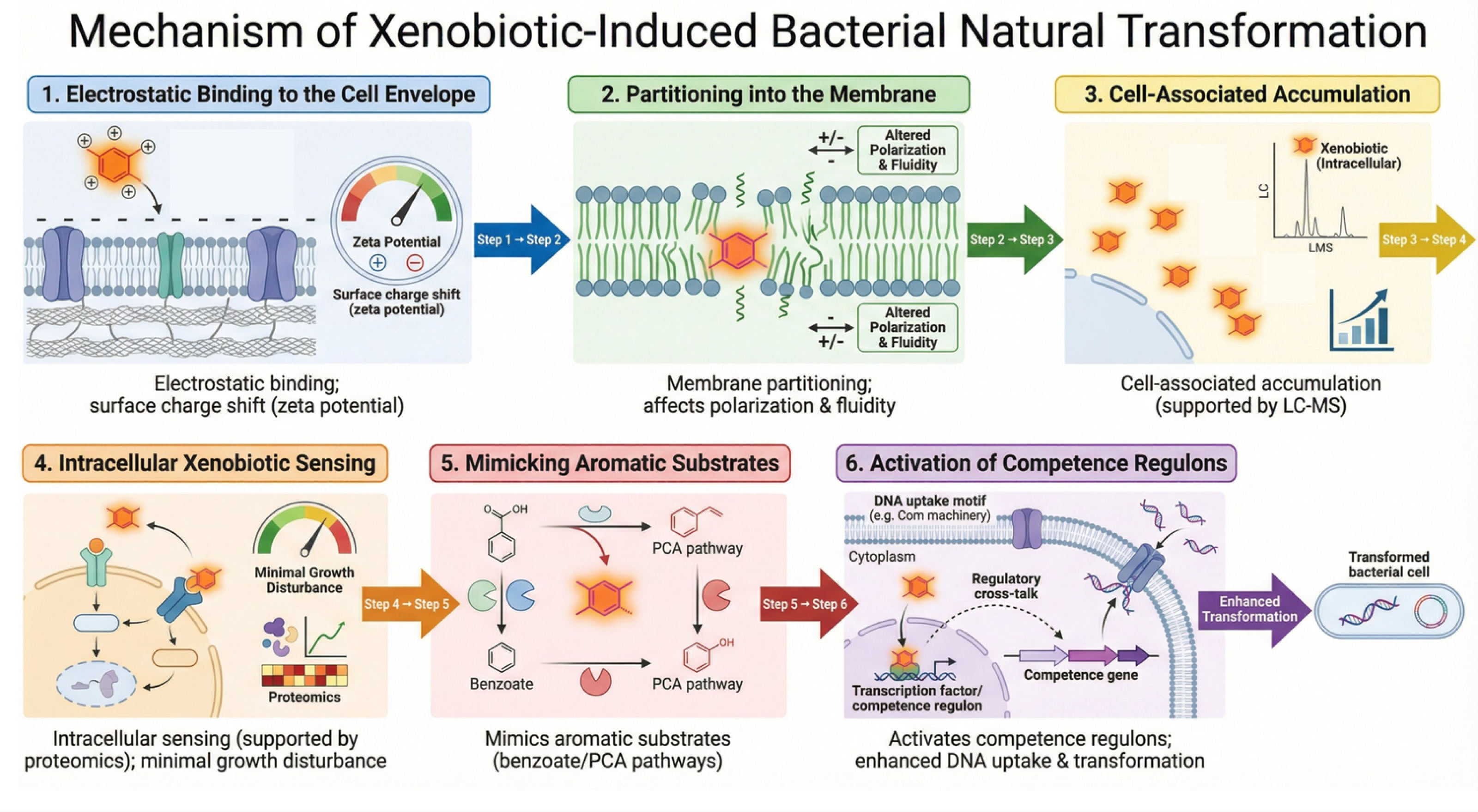
Proposed mechanism of desvenlafaxine-induced bacterial competence and transformation. This schematic illustrates the sequential steps by which desvenlafaxine interacts with bacterial cells to enhance DNA uptake and transformation: (1) Electrostatic binding to the cell envelope, inducing a surface charge shift in zeta potential; (2) Partitioning into the membrane, leading to altered polarization and fluidity; (3) Cell-associated intracellular accumulation, quantified by LC-MS; (4) Intracellular xenobiotic sensing, supported by proteomic data, with minimal growth disturbance; (5) Mimicking aromatic substrates in benzoate/PCA pathways; (6) Activation of competence regulons, resulting in enhanced DNA uptake motifs, regulatory cross-talk, transcription factor activation, and ultimately increased transformation efficiency in bacterial cells.

Our findings have important environmental implications. Because of its high water solubility and moderate lipophilicity (log P 2.75 against 3.15 for the parent venlafaxine), desvenlafaxine is weakly attenuated during conventional wastewater treatment. The molecule remains mostly in the dissolved phase and enters receiving rivers in higher proportions because the demethylation of the methoxy group to a phenolic –OH makes it much more hydrophilic and less likely to sorption onto sludge or bioaccumulation in biota (16). Strong hydrogen-bonding interactions with environmental matrices (organic matter, clays, biofilms) are made possible structurally by the free phenolic –OH, the tertiary amine, and the cyclohexanol –OH. This allows for both high mobility and remarkable persistence over long distances and depths in surface waters (51). Even though the effective concentrations in our lab assays (mg/L) exceed typical ambient levels (ng/L–µg/L), hotspots or chronic exposure scenarios (e.g. biofilms in effluent channels) might reach stimulatory levels. These results add desvenlafaxine to the growing list of pharmaceuticals that may influence antibiotic resistance via HGT. They underscore the need for One Health perspectives on drug pollution: pharmaceuticals intended for human use can have unanticipated ecological consequences.

Here we used a model laboratory strain (*A. baylyi* ADP1) and plasmid under controlled conditions. But, natural communities may respond differently, and antibiotics, mixtures of other pharmaceuticals, and nutrient levels vary. Future work should therefore (i) test long-term exposure to determine whether chronic desvenlafaxine selects for stable genetic adaptations (e.g., mutations linked to resistance/tolerance), as has been observed for antidepressant exposures in other systems (18), (ii) evaluate mixtures and co-stressors (e.g., pharmaceuticals with metals/disinfectants/UV) because combined stressors can produce non-additive transformation outcomes (13), and (iii) extend to more complex host-associated settings such as the gut microbiota, where antidepressants have been shown to reshape plasmid transfer patterns and ARG exchange among taxa, with potential downstream relevance to pathogen acquisition (19). Finally, it would also be valuable to examine the fate of acquired resistance in downstream pathogens or to explore whether similar membrane effects occur in other competent bacteria.

## Conclusion

In summary, this study demonstrates that desvenlafaxine, one of the most abundant antidepressants detected in WWTP effluents in western New York, increased natural transformation frequency in *Acinetobacter baylyi* ADP1 at mid-to-high concentrations. This effect was associated with dose-dependent increases in membrane fluidity and shifts to less negative surface charge, without evidence of ROS induction or cytotoxicity. Proteomic analysis revealed upregulation of competence-associated proteins and a coordinated metabolic response at 10 mg/L, while electron microscopy confirmed increased pili abundance at both doses. Together, these observations suggest that desvenlafaxine promotes bacterial competence through membrane-mediated mechanisms rather than stress-induced pathways. Future work should assess whether these effects extend to environmentally relevant concentrations, other bacterial species, and complex microbial communities such as those found in wastewater environments.

## Acknowledgements

This work was supported by the U.S. National Science Foundation under the Using Rules of Life to Address Societal Challenges (URoL:ASC) program, Award No. 2319520 (University at Buffalo) and 2319521 (Iowa State University). Any opinions, findings, and conclusions or recommendations expressed in this material are those of the authors and do not necessarily reflect the views of the National Science Foundation.

## Data availability

All data are available in GitHub through Zenodo (https://doi.org/10.5281/zenodo.20111088) The proteomics data generated in this study have been deposited in the ProteomeXchange Consortium via PRIDE under the dataset identifier PXD077476.

## Authors’ contribution

Funding Acquisition: DA, AH, and LJ; Study planning: NS, AH, and LJ; Study plan validation: All authors; Data collection: NS, LMA, and DA; Data analysis: NS and BR; Data interpretation: NS, AH, and LJ; First drafting: NS; Review and re-drafting: NS and LJ; Final approval: All authors.

## References

1. Rizzo L, Manaia C, Merlin C, Schwartz T, Dagot C, Ploy MC, Michael I, Fatta-Kassinos D. 2013. Urban wastewater treatment plants as hotspots for antibiotic resistant bacteria and genes spread into the environment: a review. Sci Total Environ 447:345–360.

2. Karkman A, Do TT, Walsh F, Virta MPJ. 2018. Antibiotic-Resistance Genes in Waste Water. Trends in Microbiology 26:220–228.

3. Johnston C, Martin B, Fichant G, Polard P, Claverys J-P. 2014. Bacterial transformation: distribution, shared mechanisms and divergent control. Nat Rev Microbiol 12:181–196.

4. Marutescu LG, Popa M, Gheorghe-Barbu I, Barbu IC, Rodríguez-Molina D, Berglund F, Blaak H, Flach C-F, Kemper MA, Spießberger B, Wengenroth L, Larsson DGJ, Nowak D, Radon K, de Roda Husman AM, Wieser A, Schmitt H, Pircalabioru Gradisteanu G, Vrancianu CO, Chifiriuc MC. 2023. Wastewater treatment plants, an “escape gate” for ESCAPE pathogens. Front Microbiol 14:1193907.

5. Niu C, Wu H, Wang X, Hu L, Han Y, Qiao J. 2025. Molecular mechanisms and applications of natural transformation in bacteria. Front Microbiol 16:1578813.

6. Vesel N, Stare E, Štefanič P, Floccari VA, Mandic-Mulec I, Dragoš A. 2025. Naturally competent bacteria and their genetic parasites—a battle for control over horizontal gene transfer? FEMS Microbiol Rev 49:fuaf035.

7. Rayan RA. 2023. Pharmaceutical effluent evokes superbugs in the environment: A call to action. Biosafety and Health 5:363–371.

8. Savin M, Hammerl JA, Hassa J, Hembach N, Kalinowski J, Schwartz T, Droop F, Mutters NT. 2023. Free-floating extracellular DNA (exDNA) in different wastewaters: Status quo on exDNA-associated antimicrobial resistance genes. Environmental Pollution 337:122560.

9. Matesun J, Petrik L, Musvoto E, Ayinde W, Ikumi D. 2024. Limitations of wastewater treatment plants in removing trace anthropogenic biomarkers and future directions: A review. Ecotoxicology and Environmental Safety 281:116610.

10. Zhang S, Wang Y, Song H, Lu J, Yuan Z, Guo J. 2019. Copper nanoparticles and copper ions promote horizontal transfer of plasmid-mediated multi-antibiotic resistance genes across bacterial genera. Environment International 129:478–487.

11. Wang Y, Lu J, Engelstädter J, Zhang S, Ding P, Mao L, Yuan Z, Bond PL, Guo J. 2020. Non-antibiotic pharmaceuticals enhance the transmission of exogenous antibiotic resistance genes through bacterial transformation. The ISME Journal 14:2179–2196.

12. Yu Z, Wang Y, Henderson IR, Guo J. 2022. Artificial sweeteners stimulate horizontal transfer of extracellular antibiotic resistance genes through natural transformation. The ISME Journal 16:543–554.

13. Al-Gashgari B, Mantilla-Calderon D, Wang T, De Los Angeles Gomez M, Baasher F, Daffonchio D, Laleg-Kirati T-M, Hong P-Y. 2023. Impact of chemicals and physical stressors on horizontal gene transfer via natural transformation. Nat Water 1:635–648.

14. Jia Y, Wang Z, Zhu S, Wang Z, Liu Y. 2023. Disinfectants facilitate the transformation of exogenous antibiotic resistance genes via multiple pathways. Ecotoxicology and Environmental Safety 253:114678.

15. Alav I, Buckner MMC. 2024. Non-antibiotic compounds associated with humans and the environment can promote horizontal transfer of antimicrobial resistance genes. Critical Reviews in Microbiology 50:993–1010.

16. Liu Y, Lv J, Guo C, Jin X, Zuo D, Xu J. 2025. Environmental behavior, risks, and management of antidepressants in the aquatic environment 10.1039/D4EM00793J.

17. Lu J, Ding P, Wang Y, Guo J. 2022. Antidepressants promote the spread of extracellular antibiotic resistance genes via transformation. ISME Commun 2:63.

18. Wang Y, Yu Z, Ding P, Lu J, Mao L, Ngiam L, Yuan Z, Engelstädter J, Schembri MA, Guo J. 2023. Antidepressants can induce mutation and enhance persistence toward multiple antibiotics. Proc Natl Acad Sci U S A 120:e2208344120.

19. Ding P, Lu J, Lei T, Guo Y, Zhu B, Zhao Y, Wang Y, Engelstädter J, Schembri MA, Guo J. 2025. Antidepressant drugs promote the spread of broad-host-range plasmid in mouse and human gut microbiota. Gut Microbes 17:2514138.

20. Gasser G, Pankratov I, Elhanany S, Werner P, Gun J, Gelman F, Lev O. 2012. Field and laboratory studies of the fate and enantiomeric enrichment of venlafaxine and O-desmethylvenlafaxine under aerobic and anaerobic conditions. Chemosphere 88:98–105.

21. Rúa-Gómez PC, Püttmann W. 2013. Degradation of lidocaine, tramadol, venlafaxine and the metabolites *O*-desmethyltramadol and *O*-desmethylvenlafaxine in surface waters. Chemosphere 90:1952–1959.

22. Arnnok P, Singh RR, Burakham R, Pérez-Fuentetaja A, Aga DS. 2017. Selective Uptake and Bioaccumulation of Antidepressants in Fish from Effluent-Impacted Niagara River. Environ Sci Technol 51:10652–10662.

23. Pappas JJ, DesRochers N, Tuteja B, Hughes D, McLaughlin A, Sabourin L, Renaud JB, Littlejohn C, Parrott J, Lapen DR, Sumarah MW. 2025. Ecotoxicological implications of increased antidepressant concentrations in the Laurentian Great Lakes Basin, 2018–2023. Science of The Total Environment 981:179331.

24. Gomez Cortes, Livia, Marinov, Dimitar, Sanseverino, Isabella, Navarro Cuenca, Anna, Niegowska, Magdalena, Porcel Rodriguez, Elena, Lettieri, Teresa. 2020. Selection of substances for the 3rd Watch List under the Water Framework Directive. Publications Office of the European Union. https://data.europa.eu/doi/10.2760/194067. Retrieved 13 January 2026.

25. Gomez CL, Marinov D, Sanseverino I, Navarro CA, Niegowska CM, Porcel RE, Stefanelli F, Lettieri T. 2022. Selection of substances for the 4th Watch List under the Water Framework Directive. JRC Publications Repository. https://publications.jrc.ec.europa.eu/repository/handle/JRC130252. Retrieved 13 January 2026.

26. Perry R, Cassagnol M. 2009. Desvenlafaxine: A new serotonin-norepinephrine reuptake inhibitor for the treatment of adults with major depressive disorder. Clinical Therapeutics 31:1374–1404.

27. Souza LP de, Sanches-Neto FO, Junior GMY, Ramos B, Lastre-Acosta AM, Carvalho-Silva VH, Teixeira ACSC. 2022. Photochemical environmental persistence of venlafaxine in an urban water reservoir: A combined experimental and computational investigation. Process Safety and Environmental Protection 166:478–490.

28. Young DM, Parke D, Ornston LN. 2005. Opportunities for genetic investigation afforded by Acinetobacter baylyi, a nutritionally versatile bacterial species that is highly competent for natural transformation. Annu Rev Microbiol 59:519–551.

29. Santala S, Santala V. 2021. Acinetobacter baylyi ADP1—naturally competent for synthetic biology. Essays Biochem 65:309–318.

30. Singh RR, Angeles LF, Butryn DM, Metch JW, Garner E, Vikesland PJ, Aga DS. 2019. Towards a harmonized method for the global reconnaissance of multi-class antimicrobials and other pharmaceuticals in wastewater and receiving surface waters. Environment International 124:361–369.

31. Halwatura LM, Aga DS. 2023. Broad-range extraction of highly polar to non-polar organic contaminants for inclusive target analysis and suspect screening of environmental samples. Science of The Total Environment 893:164707.

32. Guardabassi L, Petersen A, Olsen JE, Dalsgaard A. 1998. Antibiotic Resistance in Acinetobacter spp. Isolated from Sewers Receiving Waste Effluent from a Hospital and a Pharmaceutical Plant. Appl Environ Microbiol 64:3499–3502.

33. Zhang Y, Marrs CF, Simon C, Xi C. 2009. Wastewater treatment contributes to selective increase of antibiotic resistance among *Acinetobacter* spp. Science of The Total Environment 407:3702–3706.

34. Zhang S, Lu J, Wang Y, Verstraete W, Yuan Z, Guo J. 2022. Insights of metallic nanoparticles and ions in accelerating the bacterial uptake of antibiotic resistance genes. Journal of Hazardous Materials 421:126728.

35. Santoscoy MC, Jarboe LR. 2021. A systematic framework for using membrane metrics for strain engineering. Metabolic Engineering 66:98–113.

36. Pandey BN, Mishra KP. 1999. Radiation induced oxidative damage modification by cholesterol in liposomal membrane. Radiation Physics and Chemistry 54:481–489.

37. Perez-Riverol Y, Bandla C, Kundu DJ, Kamatchinathan S, Bai J, Hewapathirana S, John NS, Prakash A, Walzer M, Wang S, Vizcaíno JA. 2025. The PRIDE database at 20 years: 2025 update. Nucleic Acids Res 53:D543–D553.

38. Bernot MJ, Becker JC, Doll J, Lauer TE. 2016. A national reconnaissance of trace organic compounds (TOCs) in United States lotic ecosystems. Science of The Total Environment 572:422–433.

39. Ahmed HR, Ghafoor DD, Agha NNM, Muhamad GA, Husamadin P, Ali TM. 2025. Advanced strategies for the removal of venlafaxine from aqueous environments: a critical review of adsorption and advanced oxidation pathways. RSC Adv 15:38889–38905.

40. Ding P, Lu J, Wang Y, Schembri MA, Guo J. 2022. Antidepressants promote the spread of antibiotic resistance via horizontally conjugative gene transfer. Environ Microbiol 24:5261–5276.

41. Halder S, Yadav KK, Sarkar R, Mukherjee S, Saha P, Haldar S, Karmakar S, Sen T. 2015. Alteration of Zeta potential and membrane permeability in bacteria: a study with cationic agents. Springerplus 4:672.

42. Soon RL, Nation RL, Cockram S, Moffatt JH, Harper M, Adler B, Boyce JD, Larson I, Li J. 2011. Different surface charge of colistin-susceptible and -resistant Acinetobacter baumannii cells measured with zeta potential as a function of growth phase and colistin treatment. J Antimicrob Chemother 66:126–133.

43. Endo TH, Santos MH de M, Scandorieiro S, Gonçalves BC, Vespero EC, Perugini MRE, Pavanelli WR, Nakazato G, Kobayashi RKT. 2025. Selective Serotonin Reuptake Inhibitors: Antimicrobial Activity Against ESKAPEE Bacteria and Mechanisms of Action. Antibiotics 14.

44. Ellison CK, Dalia TN, Klancher CA, Shaevitz JW, Gitai Z, Dalia AB. 2021. Acinetobacter baylyi regulates type IV pilus synthesis by employing two extension motors and a motor protein inhibitor. Nat Commun 12:3744.

45. Ellison CK, Dalia TN, Vidal Ceballos A, Wang JC-Y, Biais N, Brun YV, Dalia AB. 2018. Retraction of DNA-bound type IV competence pili initiates DNA uptake during natural transformation in Vibrio cholerae. Nat Microbiol 3:773–780.

46. Ezezika OC, Collier-Hyams LS, Dale HA, Burk AC, Neidle EL. 2006. CatM Regulation of the benABCDE Operon: Functional Divergence of Two LysR-Type Paralogs in Acinetobacter baylyi ADP1. Appl Environ Microbiol 72:1749–1758.

47. Siehler SY, Dal S, Fischer R, Patz P, Gerischer U. 2007. Multiple-Level Regulation of Genes for Protocatechuate Degradation in Acinetobacter baylyi Includes Cross-Regulation. Appl Environ Microbiol 73:232–242.

48. Qin J, Qi X, Li Y, Tang Z, Zhang X, Ru S, Xiong J-Q. 2024. Bisphenols can promote antibiotic resistance by inducing metabolic adaptations and natural transformation. Journal of Hazardous Materials 470:134149.

49. Vergalli J, Atzori A, Pajovic J, Dumont E, Malloci G, Masi M, Vargiu AV, Winterhalter M, Réfrégiers M, Ruggerone P, Pagès J-M. 2020. The challenge of intracellular antibiotic accumulation, a function of fluoroquinolone influx versus bacterial efflux. Commun Biol 3:198.

50. Nikaido H. 2003. Molecular Basis of Bacterial Outer Membrane Permeability Revisited. Microbiol Mol Biol Rev 67:593–656.

51. Castillo-Zacarías C, Barocio ME, Hidalgo-Vázquez E, Sosa-Hernández JE, Parra-Arroyo L, López-Pacheco IY, Barceló D, Iqbal HNM, Parra-Saldívar R. 2021. Antidepressant drugs as emerging contaminants: Occurrence in urban and non-urban waters and analytical methods for their detection. Sci Total Environ 757:143722.

