## Supplemental Information for "Antidepressant desvenlafaxine identified in wastewater promotes transformation and antibiotic resistance risk in *Acinetobacter baylyi* via metabolic adaptations"

**Laura Jarboe**

Department of Chemical and Biological Engineering,

Iowa State University, Ames, IA 50011, United States.

**Supplemental Methods**

**M1. Instrumentation and LC-MS conditions**

The solid-phase extraction was performed by connecting two cartridges with different stationary phases. The Hydrophilic-Lipophilic Balance (HLB) cartridge was placed on top of a mixed-mode cation exchange (MCX) cartridge. Conditioning and equilibration were carried out using 6 mL of acetonitrile followed by 6 mL of NanoPureTM water. Samples were loaded under vacuum at a rate of 3-5 mL/min. Cartridges were eluted separately with 3 mL each of (i) methanol, (ii) acetonitrile, and (iii) a 1:1 v/v mixture of acetonitrile and ethyl acetate for the HLB cartridges. The MCX cartridges were eluted with the same series of solvents but with 5% ammonium hydroxide added to each. The resulting extract was evaporated under a stream of nitrogen gas and reconstituted to 1 mL with 50 μL of 1 μg/mL d3-diphenhydramine as an internal standard and the initial mobile phase, then filtered using 0.45 μm pore size nylon membrane syringe filters.

The reconstituted extracts were analyzed using Agilent 6460 triple quadrupole mass spectrometer, coupled with a 1200 HPLC system (Palo Alto, CA), equipped with an electrospray ionization (ESI) source operating in positive ion mode. Chromatographic separation was achieved on a Waters XSelect Premier CSH C18 column (150 mm × 2.1 mm i.d., 2.5 µm particle). Chromatographic separation was performed on a Waters XSelect Premier CSH C18 column (150 mm × 2.1 mm i.d., 2.5 µm particle size; Waters Corp., MA, USA). The mobile phase consisted of (A) 5 mM ammonium formate with 0.1% formic acid in NANOPure™ water, and (B) 5 mM ammonium formate with 0.1% formic acid in methanol. The gradient elution program began with 85% A and 15% B, held for 3 minutes, followed by a linear increase of B to 100% over 19 minutes. This was held for 3 minutes before returning to the initial conditions over 2 minutes and holding for an additional 14 minutes to re-equilibrate the column. The flow rate was maintained at 0.2 mL min⁻¹ throughout the analysis.

**Calculation of concentration**

The concentration of the analyte in the original sample is calculated using the isotope dilution method with a surrogate internal labeled (SIL) standard and response factor (RF) from calibration standards:

$$C_{\text{analyte}}\text{ }\text{(ng/L)}\text{=}\frac{C_{\text{SIL}}\times A_{\text{analyte}}}{A_{\text{SIL}}\times\text{RF}}$$

where

- $C_{\text{analyte}}$= concentration of the target analyte in the original sample (ng/L)
- $C_{\text{SIL}}$= concentration of the surrogate isotopically labeled internal standard added to the sample (ng/L)
- $A_{\text{analyte}}$= peak area of the native (unlabeled) analyte
- $A_{\text{SIL}}$= peak area of the surrogate isotopically labeled internal standard
- RF (Response Factor) = determined from calibration standards as:

$$\text{RF}=\frac{C_{\text{SIL, std}}\times A_{\text{analyte, std}}}{A_{\text{SIL, std}}\times C_{\text{analyte, std}}}$$

where the subscript "std" refers to values from calibration standard solutions.

**M2. Detailed growth-rate estimation and MICₓ calculations**

Growth rate–based dose–response analysis has been used previously in pharmacological and microbial systems to quantify drug effects from time-course data rather than static endpoints. We adopted an approach based on normalized growth rates relative to control, conceptually similar to growth rate (GR) metrics described in Hafner et al (Hafner et al., 2016).

Bacterial growth was monitored by measuring optical density over a 24-hour period. Raw OD values were first blank corrected by subtracting the corresponding medium blank at each time point:

$$\mathrm{OD}_{\mathrm{corr}}(t)=\mathrm{OD}_{\mathrm{raw}}(t)-\mathrm{OD}_{\mathrm{blank}}(t)$$

Corrected OD values ≤ 0 were excluded from further analysis.

Specific growth rates (μ) were estimated from exponential growth using log-transformed OD data. For each growth curve, the natural logarithm of blank-corrected OD was calculated, and the specific growth rate was defined as:

$$\mu=\frac{d\text{ }\ln(\mathrm{OD}_{\mathrm{corr}})}{dt}$$

Growth rates were estimated using a rolling-window linear regression approach. Briefly, ordinary least squares regression was performed for ln (OD) versus time across overlapping windows spanning four consecutive time points:

$$\ln(\mathrm{OD}_{i})=\beta_{0}+\mu\text{ }t_{i}+\varepsilon_{i}$$

where $t_{i}$represents time (hours) and μ (h^-1^) is the fitted slope. This procedure yielded a series of window-specific growth rates ($\mu_{w}$), and the maximum value across all windows was reported as the maximum specific growth rate for each replicate and condition:

$$\mu_{\max}=\max_{w}(\mu_{w})$$

This approach avoids the subjective section of exponential phase boundaries and is robust to variation in lag-phase duration.

For experiments including replicate cultures, growth rates were calculated independently for each replicate and summarized as mean ± standard deviation.

To compare growth across conditions, growth rates were normalized to the untreated control according to:

$$\mu_{\text{rel}}=\frac{\mu}{\mu_{0}}$$

where μ₀ represents the mean growth rate of the no-compound control. Relative growth ${(\mu}_{\text{rel}})$ was expressed either as μ/μ₀ or as percent change relative to control:

$$\%\Delta\mu=100\times\left( \frac{\mu}{\mu_{0}} - 1 \right)$$

Negative values indicate growth inhibition, whereas positive values indicate growth stimulation. Growth inhibition was additionally expressed as a signed percentage:

$$\text{Growth inhibition (\%)}=100\times\left( 1 - \frac{\mu}{\mu_{0}} \right)$$

Because growth responses were non-monotonic and exhibited hormesis at low concentrations, traditional MIC values (e.g., 50% or complete inhibition) were not always reached within the tested concentration range. Therefore, growth-rate–based MICₓ values were defined as the compound concentration at which growth inhibition exceeded x% relative to the untreated control.

For discrete MICₓ determination, MICₓ was defined as the lowest tested concentration at which inhibition ≥ x% was observed. When appropriate, interpolated MICₓ values were estimated on the inhibitory branch of the dose–response curve by linear interpolation between the two adjacent concentrations that bracketed the x% inhibition threshold. Interpolated MICₓ values are reported explicitly as estimates and accompanied by the corresponding tested concentration range.

All data processing, growth-rate calculations, and statistical analyses were performed in R (version 4.3.3) using custom scripts based on the tidyverse and zoo packages. Figures were generated using ggplot2.

**M3. Detailed proteomics sample preparation and LC-MS/MS analysis**

Total protein extracts were reduced with dithiothreitol (DTT), alkylated with iodoacetamide to modify cysteine residues, and digested overnight with trypsin/Lys-C. Digestion was terminated by addition of formic acid, and samples were dried in a SpeedVac. Peptides were desalted using C18 columns (Nest Group BioPureSPN Mini, HUM S18V) and dried again. A PRTC standard (Pierce part #88320) was spiked into each sample as an internal control for normalization. Peptides were separated by reversed-phase liquid chromatography using a Thermo Scientific Vanquish NEO HPLC system coupled to an Easy-Spray source. Separation was performed on a Thermo Scientific EASY-Spray PepMap Neo column (75 µm × 150 mm, part # ES75150PN). Mobile phase A consisted of 0.1% formic acid in water, and mobile phase B consisted of 80% acetonitrile/0.1% formic acid in water. The gradient and flow rate details are provided in the raw data files.

Raw mass spectrometry data from the sample (e.g., P1529-01_NS_AB_0-1.raw) were processed using Proteome Discoverer (PD) version 3.2.0.450 (Thermo Fisher Scientific) with a DIA-LFQ workflow template (LFQ_Astral_CHIMERYS_DIA_Uniprot_Acinetobacter_baylyi_PRTC). The workflow incorporated CHIMERYS (v3, on Ardia Server) for peptide identification in a library-free mode against a UniProt database (Uniprot_20241127.fasta, including Acinetobacter baylyi sequences and PRTC peptides for retention time calibration; total ~6,668 proteins, with decoys generated by reversal).

**Search and Identification:** Two sequential CHIMERYS searches were performed: an initial pre-scoring search (processing ~149,641 spectra, generating ~32,355 candidates) followed by a main search with recalibration (m/z shift: 5.18 ppm; collision energy offsets: -1.25 to +3.79 depending on charge; retention time window: -1.12 to +1.07 min, mean absolute error: 0.18 min). Parameters included trypsin digestion (up to 2 missed cleavages), variable modifications (e.g., methionine oxidation, cysteine carbamidomethylation), and fixed precursor/fragment mass tolerances derived from instrument settings. FDR was controlled at 1% (strict) and 5% (relaxed) using q-values and Mokapot for target-decoy competition, yielding ~96,055 high-confidence PSMs, ~22,288 peptide groups, and ~84,815 DIA precursors.

**Consensus Processing:** PSMs were grouped (e.g., into peptide sequence, isoform, and DIA precursor groups) and validated automatically to control peptide-level error rates. High-confidence filtering was applied (peptide length ≥6, PSM confidence: high, minimum 1 peptide per protein). Proteins were annotated using UniProt (aspects: biological process, cellular component, molecular function) and grouped with strict parsimony (2,435 protein groups). Quantification was performed via the Fragment Ions Quantifier node using summed abundances from all unique peptides (top 3 for protein roll-up), with no in-software normalization or scaling. Protein FDR validation targeted 1% (strict), resulting in ~2,151 high-confidence protein groups.

**Post-Processing and Normalization:** Display filters were applied to retain only high-confidence items (e.g., protein group FDR confidence = high). Abundances were exported as a tabular file. For normalization, intensities were scaled using the median abundance of PRTC peptides to account for technical variability and ensure quantitative comparability, generating the final PRTC-normalized protein abundance file. Total processing time was ~52 minutes. All steps were executed on a standard workstation with access to the Ardia cloud server for CHIMERYS.

Full instrument settings, acquisition parameters, and search results are available in the deposited raw data files (ProteomeXchange accession number PXD077476). Additional information on the workflows can be found in the Iowa State University Proteomics Facility tutorial: <https://www.biotech.iastate.edu/wp_biotech/wp-content/uploads/2023/06/542E.pdf>

**Supplemental Tables**

**Supplemental Table S1.** PCR primers

| Primer | Gene targeted | Primer sequence (5’-3’) | Target size (bp) | Annealing temperature (°C) |
| --- | --- | --- | --- | --- |
| *Amp-short-A* | *bla-TEM-1* | TCCGCCTCCATCCAGTCTA | 619 | 60 |
| *Amp-short-S* |  | TCCGTGTCGCCCTTATTCC |  |  |
| *tet-short-A* | *tetA* | CTCAACGGCCTCAACCTACTAC | 618 | 58 |
| *tet-short-S* |  | GCCTACAATCCATGCCAACC |  |  |

**Supplemental Table S2.** Thermocycling Conditions

| **PCR Recipe** | **Thermocycling Conditions** |
| --- | --- |
| 10 µL 5X Q5 Reaction Buffer, 1 µL 10 mM dNTPs, 2.5 µL forward primer (10 µM), 2.5 µL reverse primer (10 µM), 1–10 ng template DNA, 0.5 µL Q5 Hot Start Polymerase, nuclease-free water to 50 µ | 1 cycle of 98°C for 30 sec; 35 cycles of 98°C for 10 sec, annealing temperature for 30 sec, 72°C for 20 sec; 1 cycle of 72°C for 2 min; hold at 4°C |

**Supplemental Table S3.** Descriptive statistics for effluent antidepressant concentrations (ppt) at three WWTPs in western New York (May 2024 – May 2025) (see separate Excel file)

**Supplemental Table S4**. Significantly altered proteins at 10 mg/L and 50 mg/L desvenlafaxine vs. control (see separate Excel file)

**Supplemental Table S5**. STRING protein association network cluster annotations (see separate Excel file)

**Supplemental Figures**

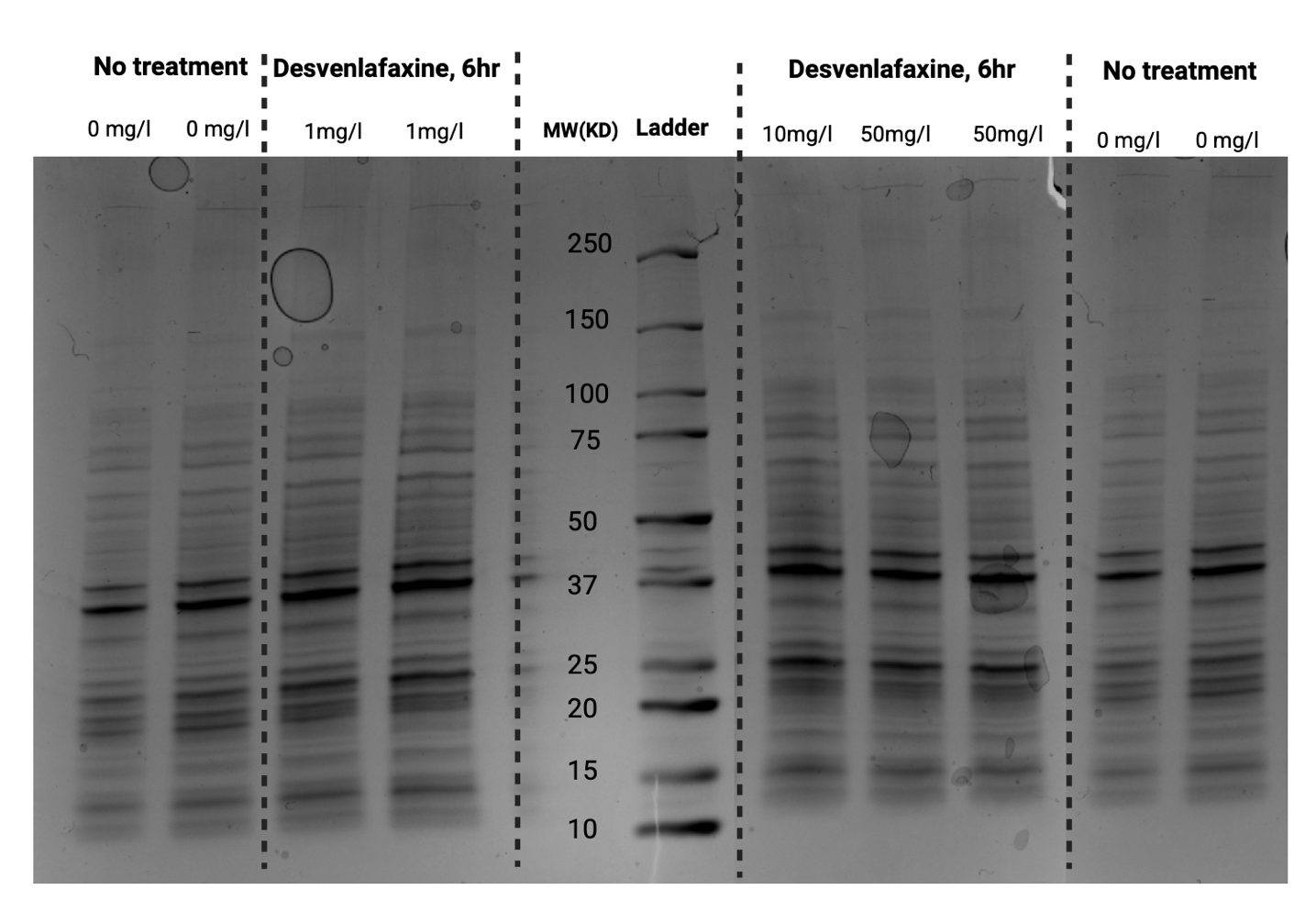

**Supplementary Figure S1.** SDS-PAGE of Desvenlafaxine induced natural transformation events. ‘No treatment’ control bands were copied to the end for easier comparison.

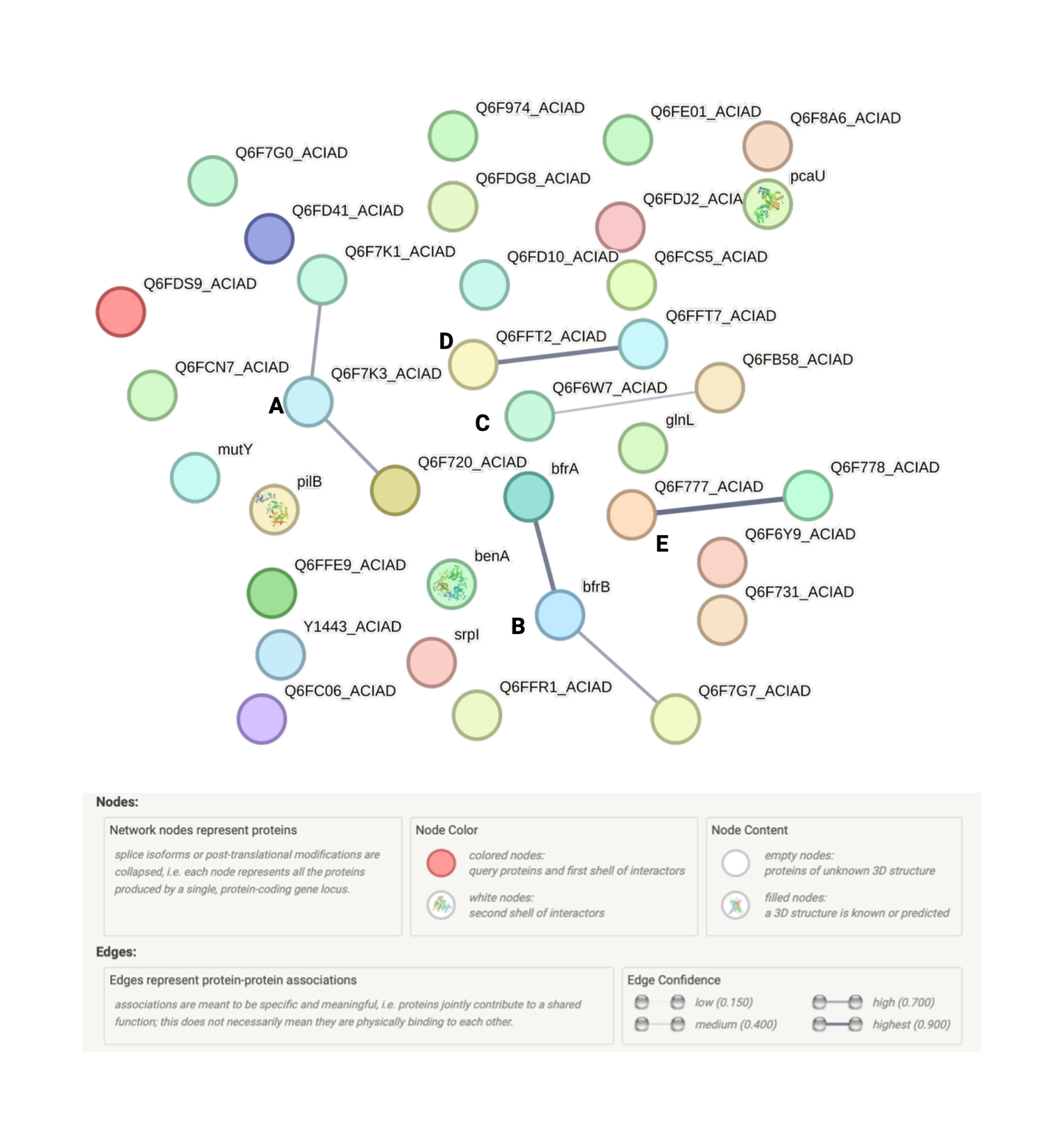
**Supplementary Figure S2.** **STRING analysis of protein-protein interaction networks for significantly altered proteins induced by desvenlafaxine**. To assess shared biological processes among the differentially expressed proteins, they were mapped using the STRING database (<https://string-db.org/>). No interactions were identified for proteins at 50 mg/L. However, proteins at 10 mg/L formed interactions categorized into five distinct networks: (A) Competence, (B) Stress response, (C) Transcriptional regulation, (D) Envelope remodeling, and (E) Host methylation switch. Nodes represent proteins (with splice isoforms or post-translational modifications collapsed by gene locus); colors indicate interaction shells (red for colored nodes and first shell of interactors, white for second shell); content shows empty nodes for unknown structures or filled nodes with known/predicted 3D structures. Edges denote protein-protein associations that are specific and meaningful (e.g., joint contribution to shared functions, not necessarily physical binding), with line thickness reflecting confidence levels (low: 0.150, medium: 0.400, high: 0.700, highest: 0.900).

References

Hafner, M., Niepel, M., Chung, M., and Sorger, P. K. (2016). Growth rate inhibition metrics correct for confounders in measuring sensitivity to cancer drugs. *Nat Methods* 13, 521–527. doi: 10.1038/nmeth.3853
